# Understanding *Plasmodium vivax* recurrent infections using an amplicon deep sequencing assay, PvAmpSeq, identity-by-descent and model-based classification

**DOI:** 10.1101/2025.05.26.25327775

**Authors:** Jason Rosado, Jiru Han, Thomas Obadia, Jacob Munro, Zeinabou Traore, Kael Schoffer, Jessica Brewster, Caitlin Bourke, Joseph M. Vinetz, Aimee R. Taylor, Michael White, Melanie Bahlo, Dionicia Gamboa, Ivo Mueller, Shazia Ruybal-Pesántez

## Abstract

*Plasmodium vivax* infections are characterised by recurrent bouts of blood-stage parasitaemia. Understanding the genetic relatedness of recurrences can help distinguish whether these are caused by relapse, reinfection, or recrudescence, which is critical to understand treatment efficacy and transmission dynamics. We developed PvAmpseq, an amplicon sequencing assay targeting 11 SNP-rich regions of the *P. vivax* genome. PvAmpSeq was applied to field isolates from a clinical trial in the Solomon Islands and a longitudinal observational cohort in Peru, and statistical models were applied for genetic classification of recurrences. In the Solomon Islands trial, where participants received antimalarials at baseline, half of the recurrent infections were caused by parasites with >50% relatedness to the baseline infection (identity-by-descent), with statistical models providing further classification as probable relapses and recrudescences, although with wide uncertainty. In the Peruvian cohort, half of the recurrent infections were caused by parasites with >22% relatedness to the baseline infection. PvAmpSeq provides high-resolution genotyping to characterise *P. vivax* recurrences, offering insights into transmission and treatment outcomes. We also discuss the nuances and limitations of available statistical methods for the classification of *P. vivax* genotyping data.

**Graphical abstract:** 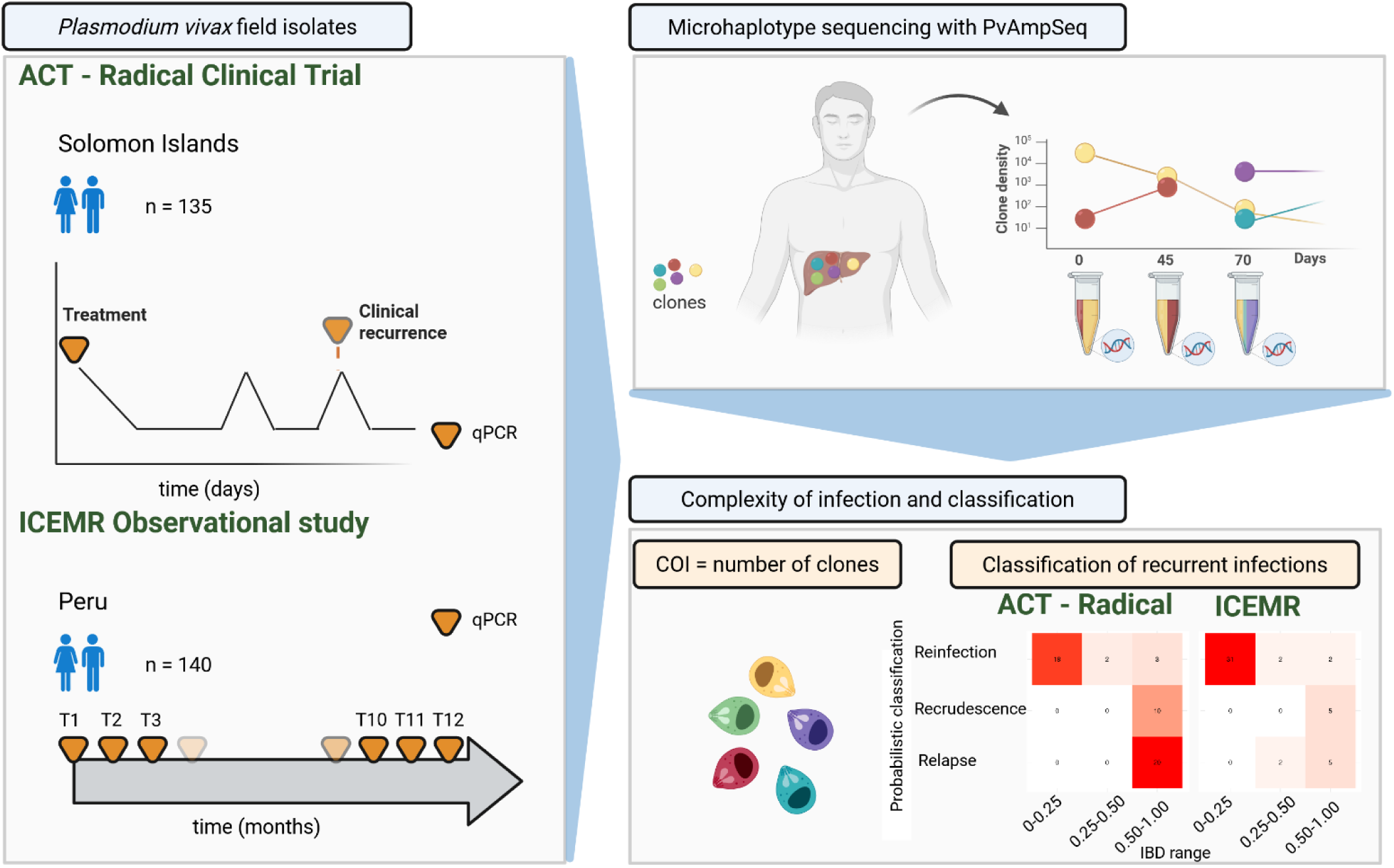

Created in BioRender. Rosado, J. (2026) https://BioRender.com/p66i686

## Introduction

*Plasmodium vivax* is the most widespread *Plasmodium* parasite infecting humans, causing an estimated 9.2 million malaria cases per year.^1^ The ability of *P. vivax* to remain in the liver as dormant hypnozoites and to cause subsequent (and often multiple) blood-stage infections contributes to onward transmission and impedes efforts to eliminate malaria.^2^ In areas endemic for *P. vivax,* recurrent *P. vivax* infections can result from relapses but also recrudescence (due to antimalarial treatment failure if the individual received treatment) or reinfections (from a new mosquito bite). Distinguishing between relapses, recrudescences, and reinfections is crucial for the assessment of the effectiveness of antimalarial-based control strategies and understanding *P. vivax* biology and epidemiology, but remains a major challenge.

The WHO Global Malaria Programme has recently highlighted the importance of using more sensitive, easily implemented, and reproducible tools that allow for estimating the efficacy of clinical drug trials.^3^ Comparing parasite genotypes of baseline infections with the ones from recurrent infections allows us to better estimate drug failure rates through ‘molecular correction’ broadly based on the identification of the same, closely related, or distinct genotypes. Unlike *P. falciparum* malaria, where genotyping must distinguish between recrudescences and reinfections, genotype data from *P. vivax* recurrent infections must be able to discriminate reinfections from both recrudescences and relapses for evaluation of treatment efficacy. Classically, genotyping infections for ‘molecular correction’ in drug-efficacy clinical trials has been done with length-polymorphic genetic markers,^4^ but more recently with amplicon sequencing approaches.^3,5,6^ A limitation of length-polymorphic genotyping can be the insufficient resolution to detect minority clones in infections.^7^ This can result in an overestimation of treatment efficacy in clinical trials. Undetected parasite clones could represent drug-resistant parasites, recrudescent clones, or hypnozoite-derived parasites, which were not detected at baseline.

The sequencing of Single Nucleotide Polymorphism (SNP) or SNP-rich amplicons by next-generation sequencing approaches (Amplicon deep sequencing or AmpSeq) has been shown to overcome the drawbacks of microsatellites.^8^ For example, i) detection of multiple SNPs in the same read allows direct detection of haplotypes, ii) sufficient sensitivity to detect minor clones and track marker-wise clone dynamics over time in *Plasmodium* infections,^7,9–11^ and iii) high reproducibility. For *P. falciparum,* AmpSeq genotyping has been shown to improve the classification of recrudescences and reinfections in clinical trials.^5,12^

In addition to their use for evaluation of treatment efficacy, AmpSeq markers are also being used to identify imported malaria cases,^13^ estimate transmission levels in population studies,^14^ detect drug-resistant parasites,^14–19^ and characterise *Plasmodium* population genetics^18,20–25^. In addition, *P. vivax* genome-wide panels of AmpSeq markers and Molecular Inversion Probes (MIPs) have recently been developed for population genetics, geographic origin assignment, and detection of SNPs associated with drug resistance (see comparative Table S1).^6,18,26–30^ In a recent study by Kleinecke and colleagues, a panel of 93 genome-wide AmpSeq microhaplotypes was applied in a clinical trial to classify 108 primary and recurrent infection pairs using identity-by-descent (IBD) estimates.^6^ The study showed a higher frequency of suspected relapses or recrudescences (high IBD) (84%) in patients not treated with primaquine (PQ) compared to those with PQ (60%).

To advance our understanding of *P. vivax* recurrent infections, we developed a *P. vivax* AmpSeq (PvAmpSeq) assay that targets 11 highly polymorphic SNP-rich regions or microhaplotypes (across 11 chromosomes) and applied several recently developed approaches for studying infections at the clone level using IBD and probabilistic models. Here, we show the application of PvAmpSeq for two cases: on samples from a clinical trial where genetic classification of recurrent infections is necessary for molecular correction of drug failure rates and from a community cohort where genetic classification is useful to understand *P. vivax* epidemiology and transmission patterns. We used samples from the artemisinin-combination therapies (ACT)-Radical randomised-controlled clinical trial conducted in the Solomon Islands between 2018 and 2019, involving patients receiving three different drug combinations, with the overall aim to test for potential antagonistic effects of the drug combinations.^31^ Two of the trial arms included drug combinations involving the anti-hypnozoite antimalarial drug PQ, where patients were treated to clear both blood-stage parasites and hypnozoites at baseline. We also used samples from an observational longitudinal cohort in the Loreto region of the Peruvian Amazon, where individuals were followed up between December 2014 and December 2015.^32^ We processed PvAmpSeq sequencing data using the AmpSeqR R package to demultiplex and analyse AmpSeq data^33^ and used various approaches for genetic classification of recurrent infections. Overall, our findings point to the potential for PvAmpSeq to provide important insights about recurrent infection dynamics in both clinical trials and epidemiological studies.

## STAR Methods

### Experimental Model and Subject Details

#### Ethics

Ethical approval for the Solomon Islands ACT-Radical clinical trial was obtained from the Solomon Islands Health Research and Ethics Review Board (HRE 041-16) and the Walter and Eliza Hall Institute of Medical Research Human Research Ethics Committee (WEHI 16-08). The Peruvian cohort study was approved by the Institutional Ethics Committee at the Universidad Peruana Cayetano Heredia (UPCH) (SIDISI 57395/2013) and by the University of California San Diego Human Subjects Protection Program (Project # 100765). UPCH also approved the use of the *P. vivax* DNA samples at the Institut Pasteur (SIDISI 100873/2017).

#### Study sites and samples

Samples from the Solomon Islands were collected as part of the ACT-Radical clinical trial (registered under Australia and New Zealand Clinical Trials Registry ANZCTR 12617000329369) conducted between September 2017 and February 2019. Briefly, 374 individuals were enrolled in the study of whom 82% (307/374) of participants had a primary symptomatic *P. vivax* infection confirmed by qPCR and were treated with artemether-lumefantrine (AL), AL plus primaquine (AL+PQ), or dihydroartemisinin-piperaquine plus primaquine (DP+PQ). Of these, 307 individuals were actively followed for up to 168 days or until a recurrent *P. vivax* infection was confirmed by light microscopy. qPCR was performed after study completion on stored samples. 191 individuals had *P. vivax* recurrent infections confirmed by light microscopy or qPCR. Of the positive blood samples, 218 (from 91 participants) were available for PvAmpseq evaluation. Blood samples were collected in EDTA tubes for the baseline infections (n = 91), whereas recurrent infection samples were collected as dried blood spots (DBS) (n = 137).

Details on the Peruvian cohort have been previously reported.^32,34^ Briefly, a three-year-long observational cohort study was conducted in Peru from December 2012 to December 2015 in two Amazonian villages in the Loreto Region: San José de Lupuna and Cahuide as part of the Amazonian International Centre of Excellence in Malaria Research (ICEMR) study.^34^ Using home-to-home and community-based screening, volunteers ≥ 3 years old were invited to be part of this cohort. Between December 2014 and December 2015, participants were monthly sampled, and 1083 out of 7612 blood samples (14.2%) were positive for *P*. *vivax* by qPCR. Of these positive samples, 449 DNA *P. vivax* positive samples (from 176 participants) were available at Institut Pasteur for genomic studies. As these samples were originally diagnosed by SYBR Green-based qPCR methods and most of them had low parasite density infections (1.55 parasites/μL [IQR: 0.74–8.38]),^32^ we performed a diagnostic Taqman qPCR to downselect amplifiable samples. This selection resulted in 274 DNA samples from 152 participants with at least 1 qPCR positive result for *P. vivax* in the last 12 months of follow-up (1 *P. vivax* infection = 65 participants, 2 *P. vivax* infections = 56 participants, 3 *P. vivax* infections = 24 participants, 4 *P. vivax* infections = 6 participants).

All cohort participants provided written informed consent for participation in both studies. Parental written consent and assent were obtained in the case of participants <18 years in the Peruvian cohort.

### Method Details

#### DNA extraction and qPCR

In the Solomon Islands ACT-Radical clinical trial, whole-blood samples from participants at baseline were collected in EDTA tubes and conserved at −20°C until DNA extraction. Genomic DNA was extracted from 200 µL of whole blood using the Favorprep 96-well genomic DNA kit (Favorgen, cat # FADWE 96004, Taiwan) and following the manufacturer’s recommendations. PBS buffer was added to the samples with insufficient volume (< 200 µL). Dried blood spots (DBS) from participants with recurrent infections were collected onto filter paper and left to dry at room temperature. DBS samples were conserved at −20°C until DNA extraction. DBS samples were cut into 6mm diameter bloodspot discs using a hole punch. Five bloodspot discs per sample were utilised for DNA extraction using the Favorprep 96-well genomic DNA kit. Genomic DNA from whole blood and DBS was eluted in 50 µL of elution buffer and stored in 96-well plates at −30°C until their use.

In the Peruvian cohort, blood samples were collected by finger prick onto filter paper and left to dry at room temperature. DNA was isolated from DBS using the E.Z.N.A. Blood DNA Mini Kit (Omega Bio-tek, Inc., Norcross, GA, US), and molecular diagnosis was performed using the Mangold method.^35,36^

For parasite density quantification, we performed a duplex TaqMan qPCR assay combining specific primers and probes targeting the *18s rRNA* gene region of *P. falciparum* and *P. vivax* in a duplex reaction, as reported by Rosanas-Urgell *et al*., with slight modifications.^37^ The reaction was prepared in a total of 13 µL containing 1X GoTaq® Probe qPCR Master Mix (Promega, USA), 769 nM of forward and reverse primers, 384 nM of probe, 1.5 µL of Nuclease-free water, and 2 µL of DNA sample. The PCR conditions consisted of an initial denaturation at 95 °C for 2 min, followed by amplification for 45 cycles of 15 s at 95 °C and 1 min at 58 °C. The assay was run in a QuantStudio TM 5 Real-Time PCR system (Applied Biosystems, USA). The number of copies of 18s rRNA DNA/µL of DNA was determined by using a standard curve from a sevenfold serial dilution to 1:10 of a plasmid at concentrations of 1 × 10^5^ copies/µL down to 1 copy/µL in nuclease-free water. Samples with late amplification (Cq value >40 & <44; <1 copy/µL) were confirmed by an extra run. *P. falciparum* and *P. vivax* primers and probes detect 3 copies of 18s rRNA DNA per genome. This implies that a concentration of 5 copies/µL in a sample is equivalent to approximately 1-2 parasites/µL.

#### Marker selection and primer design

A panel of highly informative amplicons was designed, based on whole genome sequencing (WGS) data publicly available from the MalariaGEN *Plasmodium vivax* Genome Variation version 1.0 (PvGV) Project, accessed in June 2018.^38^ Briefly, FASTQ files were downloaded from the SRA, and SNP/indel variant calling was performed according to GATK V4 best practices against the PvP01 reference genome. Of the 354 WGS samples processed, 154 were excluded due to low coverage (< 5x median coverage), high SNP missingness (>10%), multi-clonality checks (Fws < 0.85) or being from a country with too few samples remaining (n < 15; Vietnam, n = 10; Myanmar, n = 7; Malaysia, n = 6; Indonesia, n = 4; Brazil, n = 2; Laos, n = 2; China, n = 1; India, n = 1; Madagascar, n = 1; and Sri Lanka, n = 1), leaving 203 WGS samples from 6 countries (Thailand, n = 85; Cambodia, n = 31; Peru, n = 28; Colombia, n = 25; Mexico, n = 18; and Papua New Guinea, n = 16). Genomic regions were then excluded based on the coverage distribution across remaining samples as follows: the genome was divided into 1000 bp bins, and coverage was assessed with samtools bedcov^39^ for both high mapping quality (HMQ, mapping quality >= 30) and low mapping quality (LMQ, mapQ <= 30) reads. Coverage was then normalised within samples by dividing by the sample median HMQ coverage. Genomic regions were then excluded if the median across samples was greater than 1.5 or less than 0.5 or if the median proportion of HMQ coverage was less than 0.5. This method resulted in the exclusion of the majority of the hypervariable sub-telomeric regions of PvP01. The remaining genome was then searched for all regions that contained at least 4 SNPs within 140 bp, with primer design attempted using Primer3^40^ with an optimal length of 22 bp and optimal melting temperature of 60 °C, avoiding SNPs and indels in the primer region.

#### PvAmpSeq assay

The PvAmpSeq assay amplifies 11 SNP-rich regions, or microhaplotype markers, located across 11 chromosomes. Detailed protocols are described in https://doi.org/10.5281/zenodo.17474689. Briefly, PvAmpSeq libraries were prepared after amplification using a nested PCR strategy. Due to the low parasitaemia of *P. vivax* infections frequently found in field samples, parasite genomic regions encompassing marker-specific sites were enriched by multiplex primary PCRs (pPCR) with 25 cycles of amplification. Individual secondary nested PCRs (nPCR) were performed using marker-specific primers with an overhang sequence in the 5’ end and 25 cycles, enabling multiplexing of amplicons per sample. nPCR products were purified, quantified, and normalised at 15 ng/µL using QIAGEN MinElute 96 UF PCR Purification Kit (QIAGEN, Germany) and Quant-iT PicoGreen® dsDNA kit (Thermo Fisher Scientific, USA), respectively. Normalised nPCR products were pooled and performed index PCR (iPCR) using long fusion primers with P5/P7 adapters, index and overhang sequences in the backbone, limited to 10 PCR cycles.

iPCR products were quantified and normalised at 20 ng/µL, then combined into pools of equal molarity. Long fusion primers (<200 nt) and long non-specific PCR products (>400 nt) were removed from the final library by double size exclusion using 0.55 and 0.25 volumes of NucleoMag® NGS beads, respectively (Macherey-Nagel, Germany). Each pool was normalised to 10 nM and combined into a final sequencing library. Correct amplicon sizes in library pools were confirmed by Agilent D5000 ScreenTape System (Agilent Technologies, USA). The final library was sequenced on an Illumina MiSeq platform in paired-end mode using the MiSeq reagent kit v3 (600 cycles; 2 × 300 bp) with 15% spike-in of Enterobacteria phage *PhiX* control v3 (Illumina) at the Walter and Eliza Hall Institute Genomics Core (Melbourne, Australia).

#### Analysis of assay sensitivity: Serial dilution of samples, natural and mock mixed infections

The dynamic range and limit of detection (LOD) of PvAmpSeq markers were estimated by two approaches: i) serial dilution of one clinical *P. vivax* sample at 802, 80.2, 40.1, 8.02, 4.01, and 0.8 parasites/µL, and ii) evaluating the detection of minority clones in mock mixed infections. We considered a marker successfully sequenced when it had >1000 reads per sample. We determined the last sample dilution detected by the assay as the last sample with at least 7 of 11 successfully sequenced markers (40% of missingness). Samples with fewer than 7 of 11 successfully sequenced markers were filtered out due to low-quality amplification.

To determine the LOD of minority clones of the PvAmpSeq assay, we performed the assay on both natural and mock mixed *P. vivax* infections. For this study, we define a “clone” as a unique microhaplotype detected within a sample. The LOD was established as the lower within-host frequency or the highest dilution in which a minority clone was detected, in natural or mock mixed *P. vivax* infections, respectively. As part of PvAmpSeq optimisation, we selected 6 markers (Chr03, Chr05, Chr06, Chr07, Chr08 and Chr13). To specifically measure the noise and detectability of minority clones, we then performed the assay in duplicates using the 6 markers on *P. vivax* natural infections from Papua New Guinea (n = 8) and Peru (n = 12). Two monoclonal *P. vivax* DNA isolates confirmed by WGS from the Solomon Islands trial were mixed in different proportions: 1:1, 1:10, 1:50, 1:100, 1:500, 1:1000, 10:1, 50:1, 100:1, 500:1, 1000:1 and sequenced at all 11 markers. We also evaluated the detection of minority clones by artificially creating mixed infections from sequencing data using the Biostrings^41^ and ShortRead^42^ R packages. We generated artificial amplicon datasets from a sub-selection of the raw FASTQ sequences generated in this study from 14 monoclonal *P. vivax* samples sequenced at the 11 markers. We created artificial sequence data for each marker with known MOI and clone mixture proportions by randomly selecting the sequence from two samples with 10 different mixture proportions (0.1%, 0.5%, 1%, 2%, 3%, 4%, 5%, 10%, 20%, and 50%) and 7 different read counts (100, 200, 500, 1000, 2000, 5000, and 10000). For read counts of 100, 200, and 500, the mixture proportion started at 1% because we could only extract the integer sequence from two samples. For each marker, we only artificially mixed two samples of distinct microhaplotypes and non-missing reads. Additionally, to generate more realistic amplicon datasets, we also created artificial sequence data that reflected random sampling error by utilising binomial sampling with 10 different probabilities (mixture proportions as above) and 7 different read counts (as above) and repeated this 50 times. We then randomly selected 20 combinations of two distinct samples from each marker.

#### Sequencing read analysis and haplotype calling

Sequencing reads were analysed using the R package AmpSeqR version 0.1 (https://github.com/bahlolab/AmpSeqR).^33^ Sequencing reads were demultiplexed by sample and by amplicon. The overlapping sequences of paired reads were merged, and samples with a read coverage of <1,000 reads per sample were excluded from the analysis. Index, overhang sequences, and primer sequences were removed by trimming from the forward and reverse reads. In this study, a microhaplotype was defined as an amplicon sequence variant at a given locus. The minor allele frequency (MAF) was calculated for every single nucleotide in the given dataset, and then SNPs below 0.1% in frequency were removed to exclude PCR and sequencing errors. In microhaplotypes resulting from insertion and deletion (indels), such as homopolymer regions (>3 repeated bases, based on the reference sequence, e.g., “AAAA”), the sequence was adjusted to have the reference number of repeats. Microhaplotypes with low sequence identity to the *P. vivax* P01 reference genome (<75%) or with a frequency < 1%, and chimeric and singleton reads were excluded from further analysis. A marker was considered successfully sequenced if the number of reads in a given sample was over 1000 reads and passed the criteria described above. Microhaplotype calling required a minimum of 5 reads coverage per locus, a within-host haplotype frequency of ≥ 1%, and an occurrence of this microhaplotype in ≥ 2 samples over the entire dataset. All amplicon sequencing data are available under accession numbers SAMN43387238 to SAMN43387533 at the Sequence Read Archive (SRA), and the associated BioProject ID is PRJNA1153071. Microhaplotype sequences were also deposited in the open-access Github repository: https://github.com/jrosados/PvAmpSeq.

#### Population-level and within-host diversity estimates

The expected heterozygosity (H_e_) was calculated for each locus in the given dataset as described elsewhere.^43^ The within-host microhaplotype frequency was calculated as the number of reads per microhaplotype per locus over the sum of all reads per locus in a sample. Multiplicity of infection (MOI) was calculated as the highest number of distinct microhaplotypes per locus across all loci in a given sample.

#### Reproducibility of PvAmpSeq data and comparison with available microsatellite data

As part of the optimisation of PvAmpSeq, we leveraged the availability of two sample types from the Solomon Islands ACT-Radical clinical trial: red blood cell pellets and dried blood spots from the same individual, collected at the baseline visit. We compared the microhaplotype marker coverage and MOI estimates for 17 patients with both types of samples.

In addition, five DNA samples from the Peruvian cohort had matched microsatellite genotyping data based on 16 markers published by Manrique *et al*.^44^ We also compared the PvAmpSeq MOI data with MOI estimates based on microsatellite data.

#### Classification of infections into low, medium and high IBD

We used the *dcifer* R package to calculate identity-by-descent (IBD) IBD r^ and the IBD r^_total_ (i.e., inferred shared ancestry) between polyclonal infections using a statistical framework for inference that accounts for the complexity of infection (COI) and population-level allele frequencies (in our case, microhaplotype frequencies).^45^ Briefly, we first estimated naive COI, and then population-level haplotype frequencies were adjusted for the estimated COI based on our data. *Dcifer* provides estimates of r^, the relatedness between two samples, and performs a likelihood-ratio test to test the null hypothesis that two samples are unrelated (H0: r^ or IBD = 0) at a significance level of α = 0.05, adjusted for a one-sided test. For all pairs of related samples where we reject the null hypothesis, this provides statistical confidence that two pairs of samples are significantly related. In addition, in the case of samples with COI>1 we also estimate both *M’*, which is the number of related *clone* pairs between both samples, and r^_total_, which represents the ‘total’ or overall relatedness between all clones. The method makes the simplifying assumption that there is no within-host relatedness between clones in an infection. Based on IBD estimates, infections were considered as low (IBD <0.25), medium (IBD: 0.25–0.5), and high IBD (IBD ≥0.5). We used the *ggraph* R package to generate network plots and visualize the relatedness between samples.

#### Classification of recurrences into relapse, recrudescence, and reinfections

We used the recently developed *Pv3Rs* R package, which uses a probabilistic model-based framework to compute the posterior probabilities that recurrent *P. vivax* infections are recrudescences, relapses, or reinfections based on genetic data.^46^ The model accounts for IBD of parasite clones within and between episodes within a Bayesian framework that requires as input population-level allele frequencies (in our case, microhaplotype frequencies) and prior probabilities for each recurrence classification state. The model computes the probabilities of sequences of recurrent states over recurrent episodes conditional on the genetic data on a per-participant basis and outputs the per-recurrence probabilities of each state (recrudescence, relapse, and reinfection). We report individual-level posterior probabilities for each recurrence state alongside aggregated summaries of the mean posterior probability and standard deviation across all recurrences for each state, thereby propagating uncertainty into population-level estimates. As per the recommendations for the current implementation of the model, we only ran the model on participant samples where the total COI across episodes was ≤ 8. This removed only one participant from Solomon Islands (AR-359, total COI=11) and none from Peru.

To inform our prior probabilities for recurrence states of recrudescence, relapse, or reinfection in the Solomon Islands clinical trial samples, we consider different prior probability distributions on a per-patient, per-episode basis based on whether they received PQ or not, and days since the baseline episode and receiving treatment. Given there is no reported evidence suggesting widespread resistance to ACTs in the Solomon Islands, assume the probability of recrudescence is low but not negligible and constant over time and treatment groups We specified time-dependent priors reflecting expected changes in the relative contribution of relapse and reinfection during follow-up, since Solomon Islands is still a moderate transmission setting. In addition, fast-relapsing strains like the Chesson phenotype reported in the Western Pacific, relapses can occur within <2 months, sometimes even <30 days, with multiple relapses (>3) possible. ^47,49^ Accordingly, we considered three windows of time since treatment: ≤30 days, >30 days and < 90 days, ≥90 days and specified slightly higher relapse probability in early follow-up (<30 days), with a gradual shift toward a higher probability of reinfection with increasing time since treatment. With respect to the treatment administered in each arm, the original ACT-Radical clinical trial randomized participants into three treatment arms (AL; AL-PQ; dihydroartemisinin-piperaquine+PQ [DP+PQ]),^31^ however we considered only whether the participant received PQ or not due to small sample sizes. The trial results demonstrated that both PQ arms had significantly reduced rates of recurrences compared to the AL control arm, however PQ clinical efficacy was relatively low in both the AL+PQ and DP+PQ arms (∼34-44%) so we assumed that relapses were still likely despite PQ administration.^31^ In PQ-treated participants, we specified slightly lower relapse probabilities than in AL-treated participants to reflect partial hypnozoite clearance, while still allowing for substantial relapse given the observed modest PQ efficacy in the original trial. See Supplementary Table S7 for details on the prior probabilities we used.

For the Peru cohort, which was an observational community study where participants resided in two communities (Cahuide and Lupuna), participants did not receive ‘baseline’ treatment or treatment throughout the study, meaning that untreated persistent infections were possible. However, Pv3Rs is not designed to model persistent infections and assumes recurrence states are mutually exclusive (either relapse, recrudescence or reinfection). When fit to the data from Peru, Pv3Rs is misspecified and the state of recrudescence is ill-defined. In addition, recurrence states are liable to overlap (e.g., an infectious mosquito bites a person with an untreated relapse, leading to persistent relapse plus reinfection). To work around this misspecification, we can reinterpret Pv3Rs’s recurrence state definitions as follows: Pv3Rs recrudescence = persistence without added relapse or reinfection; Pv3Rs reinfection = reinfection without persistence; Pv3Rs relapse = everything else, e.g. (relapse without persistence) OR (persistence AND reinfection) etc. Prior work from the same community cohort has shown during 12-months of follow-up (which is similar to our study period), relapses accounted for a slightly higher proportion of PCR-positive recurrences in Cahuide compared to Lupuna (48% vs 35%), however these modelled outputs were not based on genetic data.^34^ We therefore assumed most recurrences in this setting are likely due to relapse or reinfection rather than recrudescence, with a slightly higher probability of relapses in participants living in Cahuide (0.45) compared to Lupuna (0.33) (Supplementary Table S8).

To estimate population-level microhaplotype frequencies, we used only the subset of baseline samples for the Solomon Islands clinical trial samples, but all samples for the Peru cohort. In the case of the Solomon Islands, we opted for this approach to minimise the potential bias from within-host selection of recrudescent parasites, which might distort microhaplotype frequency estimates if post-treatment samples were included. In the Peru cohort, we used all samples to provide a broader representation of the circulating parasite population because most recurrences are presumed to be reinfections or relapses (both drawn from the broader mosquito population).

We performed a series of sensitivity analyses to better understand the limitations of Pv3Rs and the potential impact on our results. We explored the impact of inclusion/exclusion of samples for estimation of population-level microhaplotype frequencies in the case of the Solomon Islands cohort and used all samples (baseline and follow-up) to estimate population-level microhaplotype frequencies, which did not impact the classification of samples. We also explored the impact of uniform and non-uniform priors on recurrence classification state. We did this in two ways: by skewing the prior probability completely to one classification state (e.g. prior probability of reinfection = 0.95; prior probability of recrudescence = 0.025; prior probability of relapse = 0.025, for all combinations) as well as a uniform prior assuming equal probabilities (prior probabilities of reinfection = 0.33; recrudescence = 0.33; relapse = 0.33) (Supplementary table S9).

To estimate the false discovery rate (FDR) of relapses and recrudescences (i.e., non-reinfections), we performed a simple perturbation of infection pairs. We randomised infection pairs (i.e., selecting entire ‘infection sets’ of microhaplotypes and randomly pairing them to another infection set), ensuring that pairs were not derived from the same participant and imposing a time order such that the recurrent infection always occurred after the first infection. In the Solomon Islands dataset, we randomised baseline samples with follow-up samples, and in the Peru dataset, we randomised all samples. This generated a ‘null’ data set of inter-participant infection pairs where paired infections should neither represent relapses nor recrudescences. We then ran Pv3Rs on the null data set using the default uniform prior (because we had low sample sizes to appropriately randomize within specific groups, e.g. treatment arm, time windows between episodes, community) and computed the FDR as the mean posterior probability estimate of non-reinfections (1 – the probability of reinfection averaged over inter-participant pairs). We ran 100 replicates and calculated the mean FDR and 95% confidence interval across replicates for each cohort.

The expected FDR is not zero because Pv3Rs is designed to return a non-zero posterior probability of both reinfection and relapse when genetic data are consistent with recurrent parasites that are genetically unrelated to those detected previously. For each replicate, we computed the minimum FDR as one minus the upper bound on the posterior probability of reinfection averaged across inter-participant pairs. Different data sets have different minimum FDRs because different MOI vectors induce different posterior bounds; a look-up table of posterior bounds was computed previously.^46^ We then compare the mean FDR computed using posterior probabilities with the mean minimum FDR.

To assess how particular microhaplotypes might influence our posterior probability estimates of each recurrence state, e.g., due to heterozygosity or number of alleles in the population, we iteratively re-ran the Pv3Rs analysis using informative priors as described above but omitting one marker each time.

### Quantification and Statistical Analysis

#### Statistical analysis and data accessibility

Categorical variables were compared using the two-sided Fisher’s exact test or χ2 test when required. Continuous covariates were compared using two-sided Mann-Whitney, Kruskal-Wallis, or T-test when required. Relationships between parasite density and read counts were tested using the Pearson correlation coefficient, and *p-values* were adjusted by the Benjamini-Hochberg method. The success rate per marker was defined as the number of sequenced samples that met the marker criteria divided by the total number of samples with a valid number of reads (>1000 reads per sample). We used the *d_0_* distance metric, as reported by Mideo et al.,^48^ to quantify the replicability of microhaplotype frequency estimates between technical replicates. The association between clonality (monoclonal and polyclonal) and the days since preceding infection was evaluated using a two-sided Mann-Whitney test. To evaluate the association between the change of MOI across different events from the same individuals (increase, decrease or no change) and the median time since preceding infection, we used a Kruskal-Wallis test. All statistical analyses were performed using R 4.1.0 (https://www.r-project.org/). Original data, R scripts, and algorithms developed in this study are accessible in the repository https://github.com/jrosados/PvAmpSeq. The sample and patient IDs of the original databases were not known to anyone outside the research group. De-identified datasets were generated during the current study and used to make all figures available as supplementary files or tables.

## Results

### PvAmpSeq microhaplotype marker selection

Of the 354 samples processed from PvGV, 200 high-quality WGS sequences from 6 countries (Cambodia, Colombia, Mexico, Papua New Guinea, Peru, and Thailand) were searched for all regions that contained at least 4 SNPs within a window of 140 bp. All successful polymorphic regions were then ranked by expected heterozygosity within each country, with the highest mean-ranked microhaplotype being selected for each of the 14 chromosomes (Figure S1–S2). Although we excluded the majority of the hypervariable sub-telomeric regions of PvP01 genome sequence, microhaplotypes from Chr06 and Chr12 were selected but then excluded during the PCR optimisation as they were located in the proximity of hypervariable sub-telomeric pir genes. Chr04 was excluded due to the low amplification success in Solomon Islands samples. Our panel included 11 microhaplotype markers or loci across 11 chromosomes.

The final panel of PvAmpSeq microhaplotype markers comprised three loci encoding highly polymorphic surface antigens such as Merozoite Surface Protein 1 (MSP1, Chr07, PVP01_0728900), Merozoite Surface Protein 3 (MSP3.3, Chr10, PVP01_1031500) and Apical Membrane Antigen 1 (AMA1, Chr09, PVP01_0934200); proteins involved in reticulocyte invasion, Reticulocyte Binding Protein 2a (RBP2a, Chr14, PVP01_1402400); enzymes such as Protein Tyrosine Phosphatase putative (PTP2, Chr01, PVP01_0113700), Glyoxalase I-Like Protein (GILP) putative (Chr11, PVP01_1144200); pseudogenes like Lysophospholipase putative (Chr02, PVP01_0201300); and putative proteins of unknown function such as STP1 protein (Chr05, PVP01_0533300), conserved Plasmodium protein (Chr03, PVP01_0302600 and Chr13, PVP01_1346800) and Plasmodium exported protein (Chr08, PVP01_0838000). These genes contain both SNPs and indels (Text S1).

### Application to samples from two cohort studies

A total of 492 (Solomon Islands, n = 218; Peru, n = 274) samples were sequenced for the validation of the PvAmpSeq assay. 196 samples were filtered out due to a low number of reads (<1,000 reads per sample) or low identity sequences (Solomon Islands, n = 71; Peru, n = 125). Discarded samples and successful samples had a median parasite density of 14.4 parasites/μL of DNA [IQR: 5.63–39.5] and 232 parasites/μL [IQR:34.2–1240], respectively. Additionally, samples with more than 40% (5 out of 11 loci) of missingness were discarded (Peru, n = 9), as were 10 samples from the Solomon Islands trial, due to sequencing failure of their baseline infection. Negative DNA samples and negative template controls included in sequencing runs yielded <100 reads and were filtered out.

The remaining 275 samples selected for downstream analysis corresponded to 77 participants from the Solomon Islands and 93 participants from Peru. Of the Solomon Islands participants, 41 had available samples from 1 up to 6 PCR-detected recurrent infections (total 58 samples), whereas only 40 Peruvian participants had follow-up samples, corresponding to 1 or 3 recurrent infections (total 47 samples). The median age of the participants was 10.1 years [IQR: 7.42–14.4] and 36.7 [18.0–51.5] for the Solomon Islands and Peru, respectively (Table 1). There was no significant difference between the parasite density of baseline and follow-up infections for both cohorts (*p* > 0.05). As expected for the clinical trial, febrile infections were more frequent at baseline for the Solomon Islands cohort (*p* < 0.05).

**Table 1.** Epidemiological characteristics of participants and description of their infections.

| Participant characteristics | Solomon Islands<br>n = 77 | Peru<br>n = 93 |
| --- | --- | --- |
| Age, median [IQR] | 10.1 [7.42– 14.4] | 36.7 [18.0– 51.5] |
| Sex, number female (%) | 32 (41.6%) | 51 (54.8%) |
| Treatment administered | Artemether-lumefantrine (n = 21),<br>artemether-lumefantrine + primaquine (n = 25),<br>dihydroartemisinin-piperaquine + primaquine (n = 31) | Chloroquine-primaquine (n=1) |

| Infection characteristics | n = 135 | n = 140 |
| --- | --- | --- |
| Parasite density, median [IQR] |  |  |
| Baseline | 416 parasites/μL [81.7– 793]<sup>a</sup> | 341 parasites/μL [40.6– 8560]<sup>b</sup> |
| Follow-up | 151 parasites/μL [18.1– 978] | 337 parasites/μL [77.6– 1470] |
| Febrile, number (%) |  |  |
| Baseline | 38/77 (49.4%)<sup>c</sup> | 7/93 (7.5%)<sup>d</sup> |
| Follow-up | 11/58 (19.0%) | 2/47 (4.3%) |

a Statistical difference between baseline and follow-up assessed by a two-sided Mann-Whitney test, $p = 0.2515$ . b Statistical difference between baseline and follow-up assessed by a two-sided Mann-Whitney test, $p = 0.7541$ . c Statistical difference between baseline and follow-up assessed by a two-sided Fisher’s exact test, $p = 0.0003$ . d Statistical difference between baseline and follow-up assessed by a two-sided Fisher’s exact test, $p = 0.7178$ .

### Data processing and read coverage

Reads were demultiplexed and filtered using AmpSeqR.^33^ After discarding PCR artefacts, the rate of success per locus went from 0.70 to 1 (Table S2). The median success rate amongst follow-up samples was slightly lower than for baseline samples but still around 0.90 for both cohorts (Solomon Islands; baseline median: 0.99[IQR: 0.97–1.00], vs follow-up median: 0.91[0.91– 0.95], *p* < 0.05; Peru; baseline median: 0.98 [0.80-0.98], vs follow-up median: 0.89 [0.82–0.93], *p* > 0.05). The median read coverage per marker was 4309 [2051-8129] and 4752 [2535.5–7524] for the Solomon Islands and the Peruvian cohort, respectively. Chr07 had the highest median read coverage per sample in both cohorts (9774, [4672.2–9964] for the Solomon Islands and 8734 [5560.5–9857.2] for Peru), whereas Chr01 had the lowest median coverage read (1763 [960–2326] in the Solomon Islands; and 2376 [1606–4180] in Peru) (Figure 1A).

**Figure 1.**
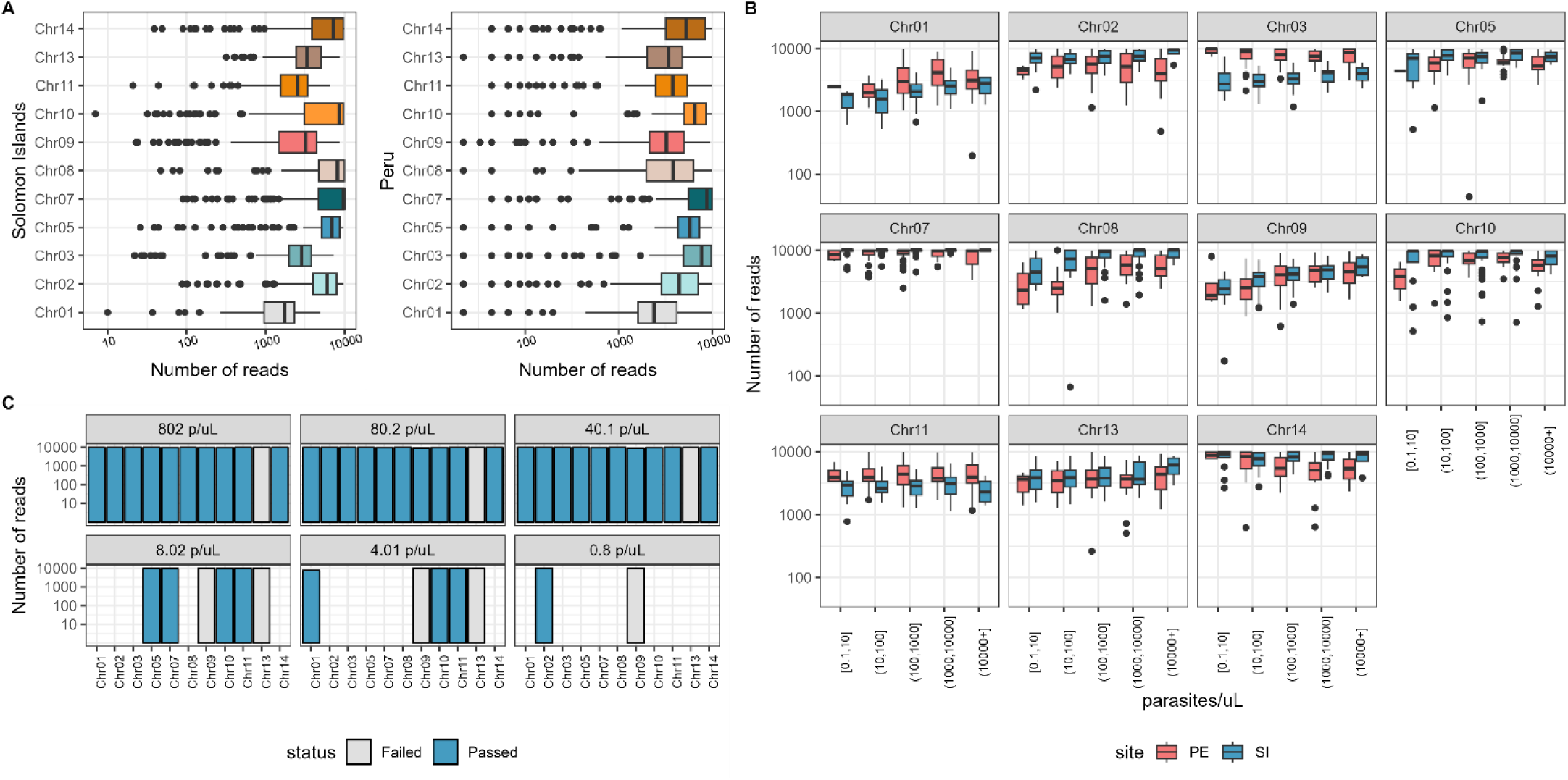
Marker coverage and sample parasite density. (A) Number of reads per marker on field samples. (B) Number of reads and parasite density of field samples. (C) Limit of detection for parasite density; in blue: markers that passed the quality control step; in grey: markers that failed the quality control step; empty bars: markers with no successful PCR amplification.

No significant correlation was found between the sample parasite density (parasites/µL) and the number of reads per marker (Pearson’s *r* range = −0.23–0.16, adjusted p range = 0.17–0.94) (Figure 1B). Samples with <10 parasites/µL were amplified with variable read coverage per marker (median: 4590 [2574–8499]). The high variance seen in each parasite density group suggested that the DNA quality affected the amplification performance. On the other hand, detection of a control sample (AR-246) was possible only when parasite density was >40 parasites/µL. Microhaplotypes were detected at dilutions of 40.1, 80.2, and 802 parasites/μL for the control sample (Figure 1C). Control sample dilutions with ≤8.02 parasites/μl were poorly sequenced for most of the loci (<60 reads). Chr13 did not successfully amplify in the control sample and was excluded from this analysis. We detected one microhaplotype per locus in the AR-246 control sample for all dilutions, except at the highest dilution of 802 parasites/μL, where two microhaplotypes were detected in marker Chr09, one of which had a very low microhaplotype frequency (0.018) (Figure S3). This microhaplotype could be a true microhaplotype since it was also detected in other samples in this dataset and with high frequency. This indicates that, as expected, higher parasite densities can more accurately determine the complexity and diversity of sample infections.

### Limit of detection for minority clones

The PvAmpSeq demonstrated high replicability and sensitivity for detecting minority *P. vivax* clones. We performed experimental replicates using 6 markers (Chr03, Chr05, Chr06, Chr07, Chr08 and Chr13) in natural infections from Papua New Guinea (n = 8) and Peru (n = 12), to measure the noise and detectability of minority clones. We found that minority clones with a within-host frequency of >1% were consistently detected in both technical duplicates providing confidence in a low LOD (Figure S4). Replicability was quantified using the *d_0_* distance metric (Mideo et al.). While minority microhaplotypes were detected across duplicates in polyclonal samples, their estimated relative within-host frequency varied between markers, and some minority clones were not detected by certain markers despite aiming for 5000 reads per marker, highlighting marker-specific performance differences in quantifying allele frequencies.

Two monoclonal samples (AR079 and AR093) from the Solomon Islands trial were selected to mimic a mixed infection. DNA samples were normalised at 100 parasites/µL and combined in the following ratios: 1:1, 1:10, 1:50, 1:100, 1:500, 1:1000, 10:1, 50:1, 100:1, 500:1, and 1000:1. The results of the mock mixed infections are shown in Figure 2A. Analyses were restricted to informative PvAmpSeq markers with distinct microhaplotypes between samples: Chr01, Chr02, Chr03, Chr05, Chr09, and Chr14. Markers Chr07, Chr08, Chr10, and Chr11 were not included because both samples had the same microhaplotype in these loci. The correct minor microhaplotype was detected in most mixtures only at 1:1 (50%), 1:10 (10%), and 10:1 (10%) mixture proportions for Chr01, Chr02, Chr09, and Chr14; however, the proportion detected often did not match the defined mixture ratio. Minority clones at 1:50 (2%) were detected for Chr02, indicating a low LOD for this particular marker. In most samples, only microhaplotypes from the major clone were detected. We examined the minor clone sequences in the raw sequence data for each sample and found that the minor clone sequence coverage was extremely low (less than 5 reads compared to over 20,000 reads in total), which resulted in the inability to detect the minor clones. These results suggest a biased PCR amplification of certain clones and a marker-specific variability in quantification. In some samples, we detected singleton microhaplotypes with very low frequencies that were not AR-079 or AR-093 microhaplotypes, which likely represent false positives (Figure S5). We compared the sequences of these major and minor microhaplotypes and found that these minor and major microhaplotypes differed in only one position. These microhaplotypes are likely to be systematic sequencing errors, with the base-call errors generally occurring at the same genomic position from different sequence reads.46 Systematic errors are often mistaken for heterozygous sites in individuals or SNPs in population analysis.

**Figure 2.**
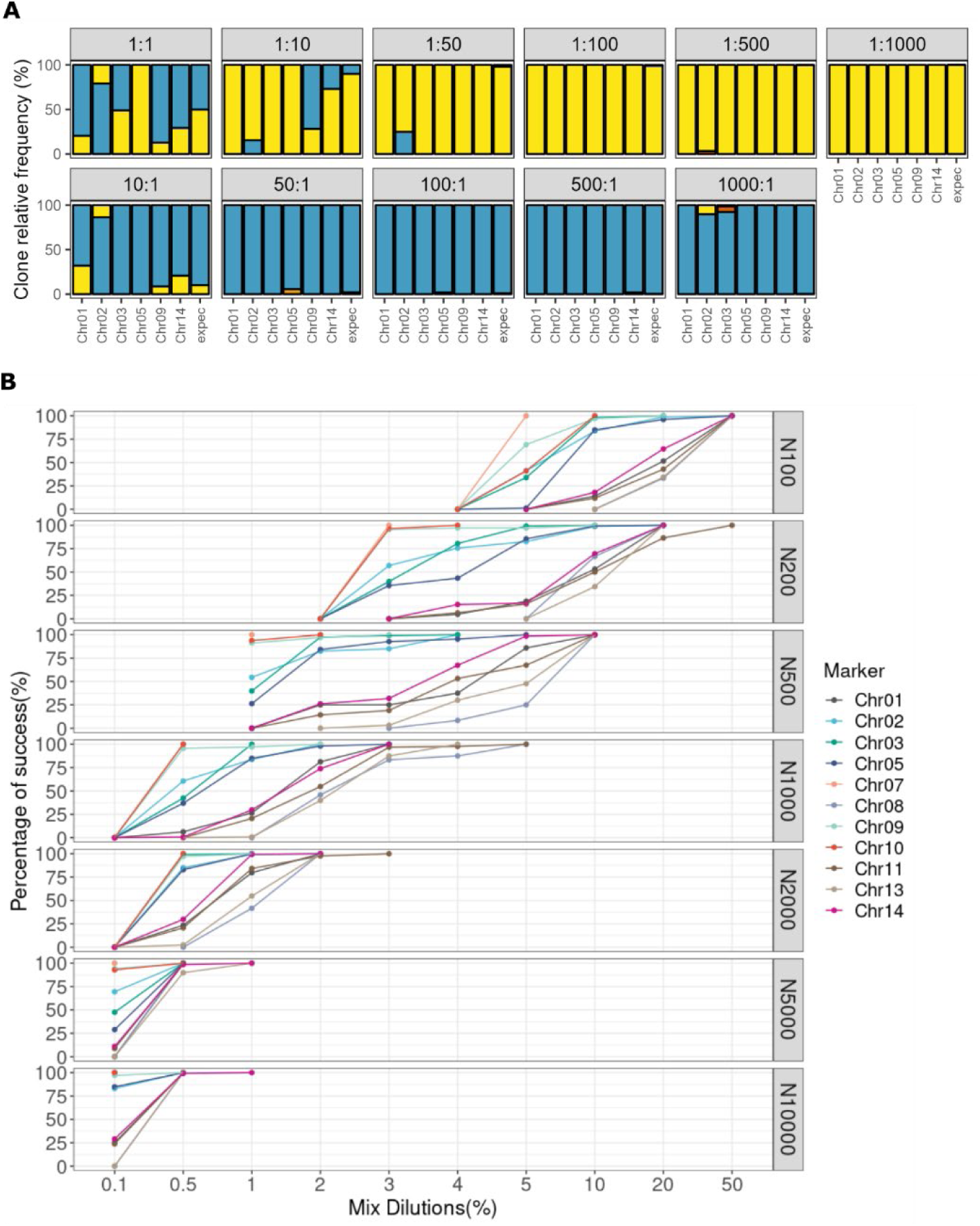
Detection of minority clones in mock and artificial mixed infections. (A) The Y-axis shows the sample ratios detected in mock mixed infections. The X-axis shows the markers grouped by expected sample ratios in mock infections (1:1, 1:10, 1:50, 1:100, 1:500, 1:1000, 10:1, 50:1, 100:1, 500:1 and 1000:1). In blue: clones from sample AR079; in yellow: clones from sample AR093; in red or orange: contaminant clones. expec: expected results from mock mixed infections. Markers sharing the same microhaplotypes between samples are not displayed. (B) The detectability of the minor clone under different numbers of reads and artificial mixture ratios. The X-axis represents the mixture dilutions (%), and the Y-axis represents the success rate (%) of detecting the minor clone. Coloured by amplicon marker.

We also examined the LOD of artificially mixed infections by determining the minority clone detectability success in silico. At the highest read counts (5,000 and 10,000), the minor clone was robustly detected at a clone relative frequency as low as 0.5% up to 50% for all amplicon markers; however, when read counts were <1,000, the minor clone was only detected accurately when the relative frequency was >10% (Figure 2B). Regardless of the data being generated with the same mixture ratios and read counts, there were substantial differences in the detectability of the minor clone based on the microhaplotype marker. For example, at a read count of 10,000 and a minor clone relative frequency of 0.1%, three markers had 100% success in detecting the minor clone (Chr03, Chr07, and Chr10). In contrast, there were very low success rates for markers Chr01, Chr08, Chr11, Chr13, and Chr14, ranging from 0% to 29%. We also created the artificially mixed infections following a binomial distribution to reflect a more realistic random sampling error and found similar results to those above (Figure S6). Combining both in vitro and in silico results of mixed infections, we thus set 2% as the lower LOD for minority clones of PvAmpSeq and recommend at least 10,000 reads per microhaplotype amplicon for robust minor clone detection (assuming frequency may be as low as 2%). If the minor clone relative frequency is expected to be around 10%, we recommend at least 1,000 reads.

### Exploring population and within-host genetic diversity metrics using PvAmpSeq

The microhaplotype markers had high diversity in both sample sets, with the mean expected heterozygosity (H_e_) of the 11 markers 0.70 for the Solomon Islands samples and 0.65 for the Peruvian samples. We found approximately half of the markers had high He ≥ 0.70 (7/11 markers in the Solomon Islands; 5/11 markers in Peru), but the markers with the highest heterozygosity differed in each cohort except for Chr05 and Chr07, which had high He in both (Figure 3A) but different microhaplotype frequencies (Figure 3B). The remaining markers had low- to moderate heterozygosity, ranging from 0.29 to 0.69 (Figure 3A). High diversity markers (He ≥ 0.70) had a median of 7 alleles (range: 4–43) and 13 microhaplotypes (range: 5–13) in the Solomon Islands and a median of 6 alleles (range: 3–34) and 7 microhaplotypes (range:5–7) in Peru (Table S3).

**Figure 3.**
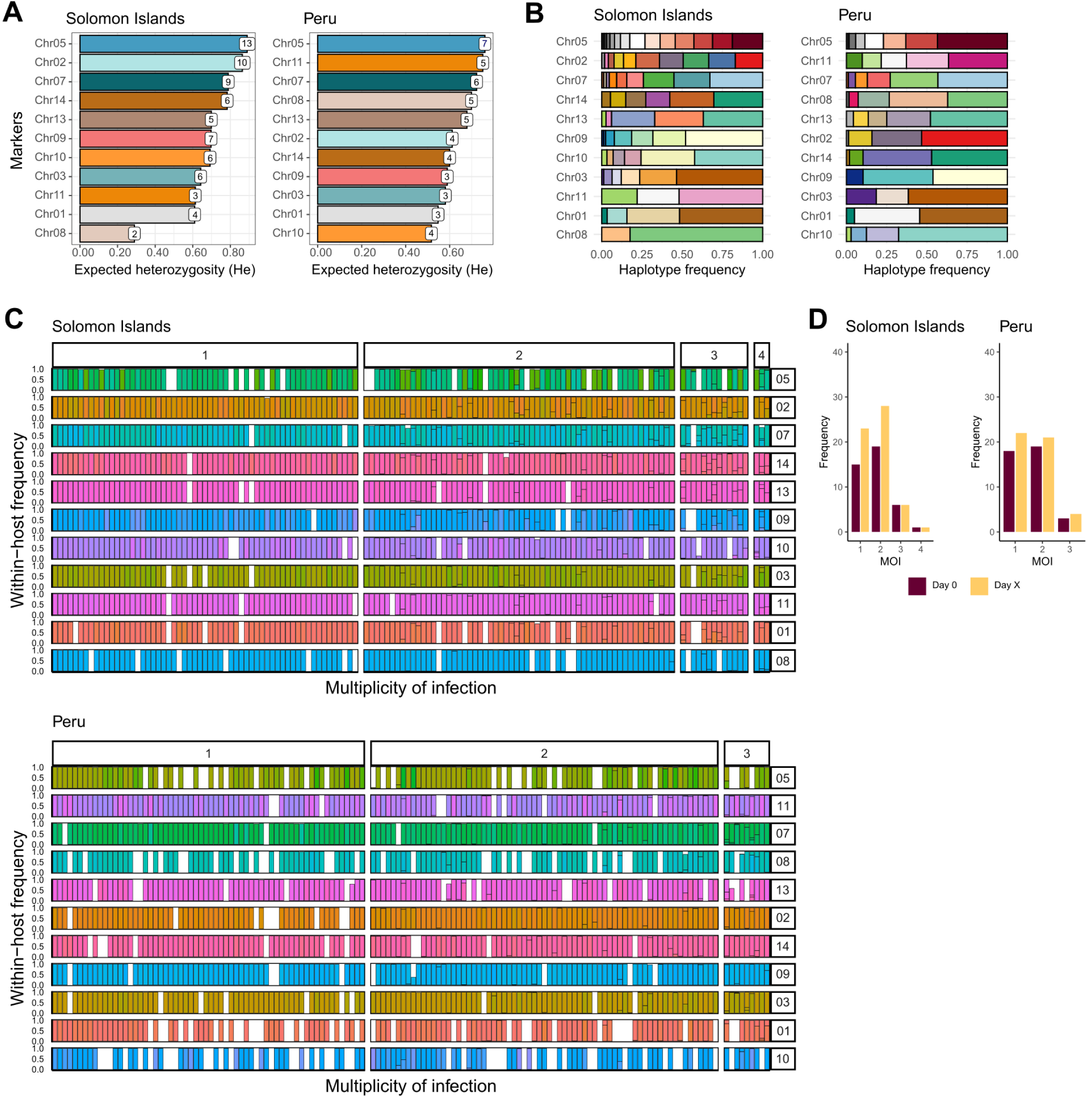
Genetic diversity of markers and multiplicity of infection (MOI) of validation samples. (A) Markers were sorted by descending H_e_. He: Expected heterozygosity calculated as the number of microhaplotypes per marker divided by microhaplotype frequency on Solomon Islands (n = 135) and Peruvian samples (n = 140). The number of microhaplotypes per marker is indicated on top of each bar. (B) Microhaplotype frequency per marker estimated for the Solomon Islands and Peruvian samples. Each colour indicates a different microhaplotype. (C) The estimated MOI in field samples as the highest number of distinct microhaplotypes by all markers. The Y-axis shows the within-host frequency of microhaplotypes per sample, calculated as the percentage of microhaplotype reads per sample. The X-axis shows the samples sorted by estimated MOI. Every sample is represented by a vertical bar. (D) Distribution of MOI in paired samples, where Day 0 represents the baseline infection (Solomon Islands, n = 41; Peru, n = 40) and Day X the day of recurrent infection (Solomon Islands, n = 58; Peru, n = 47).

Using the highest number of distinct microhaplotypes detected by all markers, we estimated the MOI in both cohorts. Out of 135 samples in the Solomon Islands cohort, 59 had a maximum MOI = 1, 60 samples had a maximum MOI = 2, 13 samples had MOI = 3, and 3 samples had MOI = 4 (Figure 3C). In the Peruvian cohort, out of 140 samples, 62 had a maximum MOI = 1, 69 samples had a maximum MOI = 2, and 9 samples had MOI = 3 (Figure 3C). No association was found between the maximum MOI obtained in baseline samples and participant’s age (Kruskal-Wallis test, Solomon Islands *p* = 0.986, Peru *p* = 0.672; Figure S7), nor with the presence of fever (χ2 test, Solomon Islands *p* = 0.744, Peru *p* = 0.823; Figure S8). There was no significant difference in MOI at baseline compared to follow-up in either cohort (Solomon Islands, baseline mean MOI = 1.83 vs follow-up mean MOI = 1.74, two-sided Mann-Whitney test, *p* = 0.5997; Peru, baseline mean MOI = 1.62 vs follow-up mean MOI = 1.62, two-sided Mann-Whitney test, *p* = 0.9244) (Figure 3D, Table S4). To test whether participants with polyclonal infections at baseline had faster times to first recurrence than those with monoclonal infections at baseline, we compared the time to first recurrent infection of paired samples stratified by MOI at baseline (MOI = 1 vs MOI ≥ 2). In both cohorts, polyclonal baseline infections (MOI ≥ 2) had comparable times to first recurrent infection to monoclonal baseline infections (MOI = 1) (two-sided Mann-Whitney test, Solomon Islands *p* = 0.7555, Peru *p* = 0.0635; Figure S9A). Likewise, the time to the next recurrent infection was not affected by the change of MOI between recurrent infections (Kruskal-Wallis test, Solomon Islands *p* = 0.3979, Peru *p* = 0.4724; Figure S9B).

As part of the assay validation, we used available microsatellite data for 5 Peruvian samples to compare MOI estimates using microsatellite genotyping and PvAmpSeq. Three out of the five samples appeared to be polyclonal by PvAmpSeq, whereas only one sample was previously reported as polyclonal by microsatellites (Table S5).

### Classifying recurrent infections using identity-by-descent and probabilistic estimates

Paired samples were further analysed to identify whether recurrent infections exhibited high or low relatedness relative to the baseline infection in the Solomon Islands trial or the first infection during the Peruvian community cohort study period. Based on IBD estimates, in the Solomon Islands ACT-Radical clinical trial, 55.2% (32/58) of recurrent infections had high IBD (IBD ≥0.5), while 41.4% (24/58) had low IBD (IBD <0.25) and 3.4% (2/58) had medium IBD (IBD 0.25–0.5) (Figure 4A). A smaller proportion of recurrences had high IBD compared to baseline among people who received primaquine at baseline, compared with people who did not, but without significant differences (68.2% [15/22] vs 47.2% [17/36]; two-sided Fisher’s exact test, *p* = 0.3202, Table S6).

**Figure 4.**
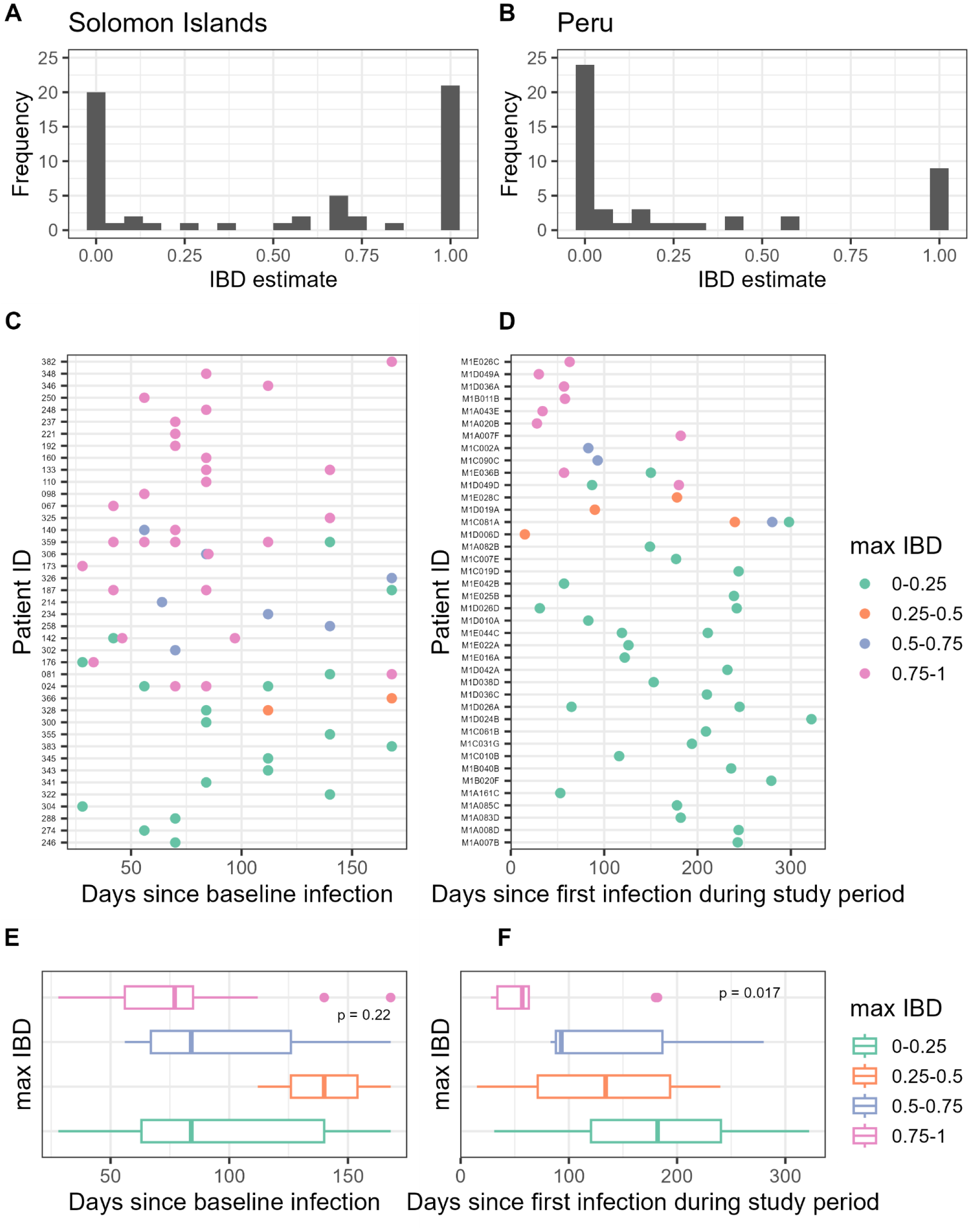
Classification of *P. vivax* recurrent infections with PvAmpSeq using IBD. (A-B). Distribution of IBD estimates in paired samples from the Solomon Islands and Peru, comparing baseline and first recurrence pairs. (C-D). Maximum IBD between recurrence and preceding infection pairs in the Solomon Islands and Peru. The X-axis shows the days since baseline (becoming infected), the Y-axis depicts each patient’s infection over time, and the colour represents the IBD estimate range for the infection pairs. (E-F). Association of time since baseline infection and maximum IBD estimate in recurrence and preceding infection pairs. Baseline (Day 0) infections are not displayed for easier interpretation of plots.

In the Peruvian community cohort, 23.4% (11/47) of recurrent infections had high IBD, while 68.1% (32/47) had low IBD and 8.5% (4/47) had medium IBD (Figure 4B). To evaluate whether recurrent polyclonal infections contribute to increased IBD, we compared the frequency of polyclonal infections in both cohorts per IBD group. We did not find an association between MOI and IBD of recurrent infections in either cohort (two-sided Fisher’s exact test, Solomon Islands *p* = 0.1279, Peru *p* = 0.3754; Table S6). Detailed microhaplotype data and IBD classification of recurrent samples can be seen in Figure S10, S11, Text S2 and Text S3.

We found that IBD in recurrent infections changed with time for most of the recurrent episodes in both cohorts, with a maximum number of five and three recurrences experienced by the same participant in the Solomon Islands and Peru, respectively. For example, we found evidence that the participants in the Solomon Islands clinical trial who had four and five recurrences had recurrences with low and high IBD, respectively, compared to the baseline infection, (Figure 4C) and in Peru, most participants experienced recurrences with low IBD (Figure 4D). There was no clear pattern with time to recurrence in the Solomon Islands clinical trial (Kruskal-Wallis test, *p* = 0.218; Figure 4E, see Figure S12A for samples that failed sequencing). In Peru, infections with high IBD tended to happen within 100 days since first infection (median 57 days [34-63]), compared to infections with low IBD that happened around a median of 188 days (range: 121-242)(Kruskal-Wallis test, *p* = 0.017; Figure 4F, see Figure S12B for samples that failed sequencing).

We evaluated the genetic relatedness (IBD estimates) of parasite isolates within and between transmission seasons in both cohorts. Higher levels of relatedness were found in the Peruvian cohort in contrast to the Solomon Islands trial (Figure S13), with possible clonal expansion (IBD ≥0.9) in the former. We detected more genetic clusters when including all samples MOI ≥1 (IBD ≥0.9), which included isolates from different transmission periods within a year and from different years.

We also performed probabilistic classification of recurrences using the Pv3Rs R package to provide further granularity on probable recurrence states. For the Solomon Islands, we explored the most probable state of each recurrent infection and found that, after accounting for uncertainty, 37.6% of participants experienced a probable reinfection (standard deviation, SD 0.41), 16.4% experienced a probable recrudescence (SD 0.30) and 46.0% experienced a potential relapse (SD 0.36) (Figure 5A, 5C). Among participants who experienced more than one recurrence, there were different sequences of most probable states, e.g. AR-024 has most probable reinfection at first recurrence followed by relapseand then a reinfection (Figure 5A). Interestingly, for this participant, the IBD estimates for recurrences were all <0.25 compared to baseline but the maximum IBD of recurrences compared to all preceding episodes were 0, 1, 1, and 0 supporting the probabilistic estimates of both reinfection and relapse (Figure 4C). A sensitivity analysis using inter-participant episode pairs generated a mean FDR of 20.6% (95% CI: 20.1–21.1%) for the Pv3Rs model. It exceeds a mean minimum FDR of 15.7% by 4.9.

**Figure 5.**
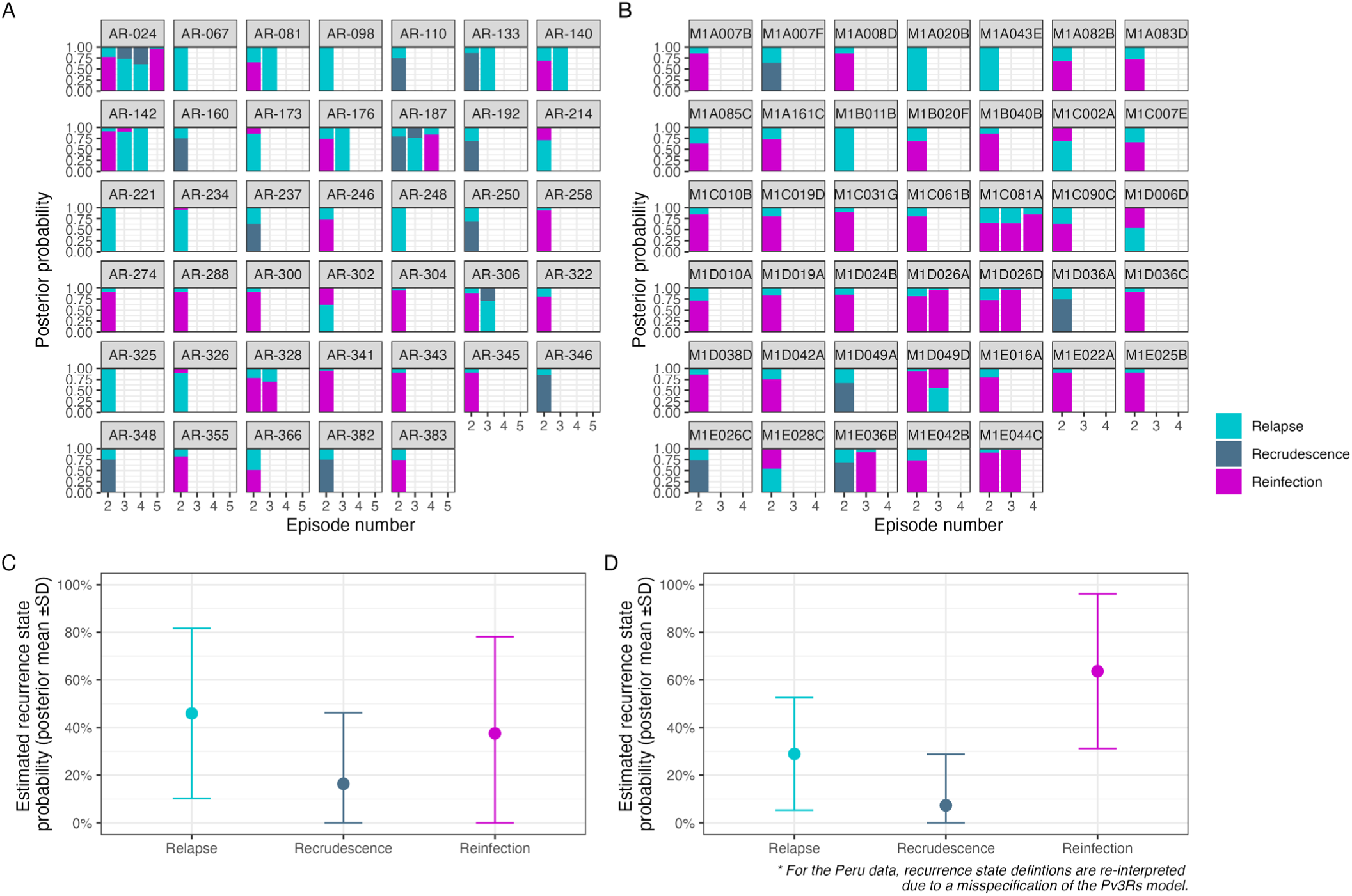
Model-based classification of recurrent *P. vivax* infections with PvAmpSeq. (A-B) Marginal posterior probabilities of relapse, recrudescence, and reinfection for each recurrent infection in each participant in (A) the Solomon Islands and (B) Peru. The X-axis shows the episode number; for example, considering the baseline infection as the first episode, the first recurrence would be episode 2. Note: this refers to successfully genotyped episodes only (*see Supplementary Figure S12*). (C-D) Mean posterior probability estimates and standard deviation for each recurrence state in (C) the Solomon Islands and (D) Peru samples. For the Peru data, due to a Pv3Rs model misspecification, recurrence state definitions are re-interpreted as follows recrudescence = persistence without added relapse or reinfection; reinfection = reinfection without persistence; relapse = everything else, e.g. (relapse without persistence) OR (persistence AND reinfection) etc (see Methods).

For Peru, after accounting for uncertainty, we found that 63.7% (SD 0.32) of participants experienced a probable reinfection without persistence of a preceding infection (Pv3Rs reinfection), 7.3% (SD 0.24) experienced a probable persistent infection without added relapse or reinfection (Pv3Rs recrudescence), and 29.0% experienced a probable relapse (SD 0.24)(Figure 5B, 5D; see Methods for details on re-interpretation of Pv3Rs recurrence state definitions). Most participants (82%, SD 0.20) experiencing more than one recurrence experienced probable reinfections without persistence (Fig 5B). A sensitivity analysis using data on inter-participant episode pairs generated a mean FDR of 27.0% (95% CI: 26.2–27.8 %) for the Pv3Rs model. It exceeds a mean minimum FDR of 16.2% by 10.8.

We explored whether the exclusion of specific markers may lead to discrepant recurrence classification. Most results, particularly for Peru, did not appear to be sensitive to genotyping errors, at least not at a single marker. In 9 of 100 recurrences (9%), removing a single marker resulted in a non-zero recrudescence probability, with no single marker driving this effect (Figure S14). Overall, we found moderate concordance between IBD estimate ranges and probabilistic classification results, with the classification of high IBD recurrences also found to be either recrudescences or relapses and a few reinfections. Similarly, low IBD recurrences were mostly associated with reinfections and a few relapses or recrudescences, which is in line with the possibility of low-IBD heterologous relapses (Figure S15A-B). This was also consistent with the microhaplotype-specific data for recurrences (Figure S16).

## Discussion

We developed PvAmpSeq, a targeted amplicon sequencing assay that enables the characterisation of *P. vivax* infections at the per-marker clonal level by targeting 11 highly polymorphic microhaplotype markers. The PvAmpSeq assay amplifies field samples with >10 parasites/µL, yielding high-depth coverage but with less coverage for samples with 1–10 parasites/µL. Both archived dried blood samples and red-blood-cell pellet samples were successfully amplified. The high coverage per marker of PvAmpSeq allows the detection of minority clones at a frequency of >2%. The PvAmpSeq microhaplotype markers comprise loci encoding surface antigens, proteins involved in reticulocyte invasion, enzymes, and proteins of unknown function. As a result of immune selection acting on these loci, their high diversity has made them suitable for measuring MOI and for tracking individual clones over the course of natural infections within an individual.^50^ Our results show that the PvAmpSeq markers provide high resolution to estimate IBD in recurrent infections. The less polymorphic PvAmpSeq markers of the panel could be suitable for studying diversity and structure between global regions; however, further exploration is needed to validate the assay with samples from various geographical origins and in comparison to other genotyping panels (e.g.,^6,26,27,29,30^).

We implemented IBD to classify recurrent infections into low (<0.25 IBD), medium (0.25-0.5 IBD) and high IBD (≥0.5 IBD) categories compared to their baseline or preceding infection(s) and using recently developed methods,^45,46^ computed the probability that recurrent infections were either relapses, recrudescences, or reinfections. At the end of the follow-up period, high IBD infections represented more than half of recurrent malarial episodes in the Solomon Islands clinical trial, which was in line with our model estimates. After accounting for uncertainty, we found that 46% of recurrences were probable relapses and 16% were probable recrudescences.

Our results are consistent with moderate levels of malaria transmission before the ACT-Radical clinical trial was conducted in the Solomon Islands,^32^ with a correspondingly high diverse parasite population. In addition, we expected more moderate-to-high transmission levels with possible parasite clonal expansion in both sampled communities of the Peruvian Amazon, Lupuna and Cahuide, based on previous work using microsatellite genotyping.^32,34,44^ Relapsing parasites with high relatedness to primary infections (high IBD) are often seen in travellers, who are likely exposed to a small number of infected mosquito bites; however, this same pattern can also be seen in individuals in endemic areas with a low hypnozoite load in their liver resulting in monoclonal relapses.^51–55^ That relapses are involved is based on the assumption that whereas some sporozoites inoculated by the mosquito become hypnozoites, others with the same genotype do not.^56^ Conversely, individuals in endemic areas are more likely to experience heterologous relapse caused by parasites with low IBD sharing due to latent hypnozoites from previous inoculations.^51,55,57^ Our data suggest that the Solomon Islands cohort had a high force of infection (FOI) and likely high hypnozoite liver burden; thus, polyclonal and low IBD relapses are expected to happen even in children. However, the weak population structure in the Solomon Islands, suggests a frequent inbreeding environment in which the inoculation of meiotic siblings from a single mosquito bite is also possible, as suggested in earlier studies.^58,59^ Similarly, low IBD recurrences relative to all previous infections were more frequent in the Peruvian cohort, suggesting they could encompass both relapses arising from hypnozoites from previous inoculations and new infections. While the low IBD recurrences in Peru align with the heterologous relapses pattern, the strong population-level connectivity makes it difficult to distinguish these from reinfections. Our results showed that recurrences whose most probable state was relapse (and could be re-interpreted as probable persistence and reinfection) were polyclonal and exhibited varying degrees of relatedness to the primary infection, supporting previous findings in Southeast Asia and Brazil. ^57,60^

We did not find a significant change of MOI over time, in contrast to findings reported by Noviyanti *et al*. ^53^, who reported decreasing MOI in subsequent relapses in Indonesian soldiers returning to a malaria-free setting, which was attributed to hypnozoite depletion in successive relapses. These discrepancies are likely due to our reliance on endemic cohorts with individuals residing in an area with moderate-to-high transmission levels, in which both relapses and new infections occur frequently, and possibly overlapping, thereby sustaining a high overall MOI. This is further supported by our finding of many probable reinfections without persistence as well as evidence of relapse and/or persisting infections in the Peru cohort.

IBD estimates (r^) correlated well with model-based classification for most recurrent infections in Peru and the Solomon Islands, showing the potential application of the PvAmpSeq panel, in particular for reinfections where we expect low IBD. In the case of “discordant” results, for instance where reinfections were found to also have medium to high IBD, we were unable to definitively ascertain whether this was due to model misspecifications/limitations (see below for discussion on model misspecifications for the Peru data) or limitations of the current PvAmpSeq panel. It is likely a combination of both since the PvAmpSeq panel includes 11 microhaplotype markers and thus may not always allow appropriate classification of recurrent *P. vivax* malaria infections. In some cases, more than 11 microhaplotype markers may be required, depending on the circulating parasite population’s genetic diversity. It is also possible that recurrence classification cannot be inferred from genetic data alone. In addition, the *dcifer*-based IBD estimates assume no within-host relatedness between clones, which would be inapplicable in the case of *P. vivax* relapses and may impact our estimates; however, simulations by Gerlovina *et al*. showed a relatively robust estimation of IBD even when this assumption is violated.^45^ Our Pv3Rs sensitivity analyses where prior probabilities were varied from uniform to skewed single-state priors showed that the impact on the posterior was low since population-level posterior means were largely preserved, indicating robustness of Pv3Rs to prior specification. In addition, relative to its minimum, the FDR for non-reinfection states (e.g. relapses and recrudescences) was small. Together these results support reporting population-level (mean) posterior probabilities, where uncertainty is propagated, rather than relying on hard or threshold-based individual-level assignments. The relative FDR was higher in Peru, in line with lower allelic diversity. Moreover, there may be potential limitations for population-based epidemiological studies (such as that in Peru) where there is no clear ‘baseline’ infection information given ongoing transmission and low treatment rates. A caveat of the current implementation of the Pv3Rs model is that Pv3Rs was designed assuming study participants are frequently and actively followed (and treated), which is appropriate for clinical trials but less so for observational studies like the Peru dataset. More specifically, Pv3Rs is not designed to model persistent infections. In studies where infections persist untreated (e.g., the Peruvian cohort), the state of recrudescence is ill-defined and recurrence states are liable to overlap (e.g., an infectious mosquito bites a person with an untreated relapse, leading to persistent relapse plus reinfection). Otherwise stated, when fit to the data from Peru, Pv3Rs is misspecified. To work around this misspecification, we can reinterpret Pv3Rs’s recurrence state definitions. In addition to true reinfections, the high rate of reinfection that we observed in Peru could be due to people having high hypnozoite burdens at enrollment and/or due to missing intermediate data (e.g. undetected recurrences/recurrences that were not successfully genotyped). Another challenge for IBD-based and Pv3Rs tools is distinguishing between recrudescence and intermittent patency in peripheral blood in chronic spleen-positive infections, i.e., a person might be continuously infected in the spleen,^61^ but parasites (all or particular clones) may only be intermittently detectable in the bloodstream; in particular, in observational studies. Future work could explore refining the model-based estimates by using time-to-event models to generate informative prior estimates, as was done with microsatellite data,^62^ and leveraging longitudinal studies with repeated sampling. Development of new models or model extensions may also be needed to robustly classify recurrent infections to account for sequencing error, given the availability of many *P. vivax* amplicon sequencing approaches, as well as explicitly considering the possibility of infection persistence (i.e., relaxing the assumption of mutually exclusive recurrence states), but this was beyond the scope of the current study. In the context of newly developed marker panels (i.e., amplicon sequencing, MIPs, and microhaplotype genotyping panels),^6,26,27,29,30^ future work could focus on rigorous benchmarking of these analysis tools to provide guidance for the community (e.g., PGEforge https://mrc-ide.github.io/PGEforge/).

Detection of microhaplotypes derived from persisting gametocytes or residual DNA in recurrent infections after ACT could overestimate treatment failure rates.^63,64^ Unlike *P. falciparum*, *P. vivax* gametocytes are commonly detected at densities of <10% of asexual parasites, and in the absence of treatment, the duration of gametocytaemia is 3 days.^65^ Our approach aimed for coverage of 10,000 reads per marker per sample, ensuring high sensitivity for detecting minority clones at frequencies of 0.5% within-host, i.e., 50 reads. Our sensitivity analysis showed we can confidently detect microhaplotypes with ≥2% within-host frequency; thus, this approach will likely detect gametocyte microhaplotypes. Interestingly, in the Solomon Islands drug-efficacy clinical trial, no recurrent infections were detected earlier than 28 days post-treatment, and high IBD recurrences were more frequent in patients treated without primaquine, suggestive of relapses; however, low sample sizes when stratifying by treatment arm limited us from drawing definitive conclusions.

A limitation of our study was the number of samples included in the validation of PvAmpSeq. The original ACT-Radical clinical trial involved 374 participants, of whom 307 completed follow-up,^31^ but the low parasite density *P. vivax* infections during follow-up limited the number of paired baseline/follow-up samples available for inclusion in PvAmpSeq validation. This could also show a potential bias of PvAmpSeq to better detect new infections that usually have high parasite density. After data demultiplexing and quality control, the remaining sample size per treatment arm was small; thus, we could not definitively employ molecular correction to conclude the treatment efficacy assessed by PvAmpSeq. Future studies should focus on enhancing the analytical sensitivity of PvAmpSeq to successfully sequence samples with low parasitaemia. This methodological improvement will enable the use of larger sample sets and paired samples to evaluate its application for informing on recurrences, as shown by Kleinecke *et al*.^6^ The sensitivity of detection of minority clones was affected by the absence of use of genetically different *P. vivax* strains for mimicking mixed infections. However, the artificial mixed infection estimated that the LOD of minority clones was 2% when we had 10,000 reads and 1% if we had 50,000 reads. This LOD is an improvement over the 10% reported by Kattenberg *et al*.,^27^ while other AmpSeq panels have not yet reported this information. With these features, the potential applications of PvAmpSeq we envision are i) the study of relapse biology in longitudinal studies, ii) the contribution of hidden parasite biomass in spleen and bone marrow to recurrent *P. vivax* infections,^66^ iii) parasite diversity dynamics from human-host to mosquito vector, iv) parasite population diversity within mosquitoes collected in cross-sectional studies.

Likewise, in the Peruvian cohort, some intermediate low-density/submicroscopic infections could have been missed; thus, some of the recurrent infections might not reflect the real infection dynamics in this population. The DNA quality from low parasite density samples represented a challenge for PvAmpSeq, as observed in samples with 1–10 parasites/µL that were discarded due to low coverage (38% of the Solomon Islands samples and 49% of the Peruvian samples). Selective Whole Genome Amplification (sWGA) has been demonstrated to improve sample coverage in very low *Plasmodium* density samples for AmpSeq or WGS methods.^16–18,27,67^ However, this type of treatment to low-density samples could incur amplification biases of the most dominant clone, increase the error rate and raise costs, making it less applicable in low-resources settings.^17,18^ Other factors may also influence the applicability of PvAmpSeq, like daily fluctuations in clone densities within infection, which may impede robust longitudinal tracking of clones, particularly minor clones. Although we tested 6 PvAmpSeq markers in paired dried-blood spot and red-blood-cell pellet samples, we were not able to evaluate the complete panel of markers in this sample subset due to limited DNA quantities. Nevertheless, we found that PvAmpSeq performed well on both sample types, albeit with better recovery of the number of microhaplotype variants for some of the markers. PvAmpSeq detected more polyclonal infections than microsatellite genotyping in a small subset of Peruvian samples; however, future studies should include larger sample sizes to evaluate the applicability of PvAmpSeq to quantify polyclonal infections in other settings.

AmpSeq has been proposed as a new gold standard for analysing malaria drug clinical trials.^3,12^ For *P. falciparum*, five AmpSeq markers have been validated to discern between recrudescences and reinfections.^5^ Regardless of the level of endemicity and the cut-off of detection for minority clones (0.1–2%), simulation studies showed that the use of 3 to 5 polymorphic markers was sufficient to classify *P. falciparum* recurrent infections as recrudescences or reinfections.^12^ In *P. vivax* infections, genotyping of highly related sibling progeny present in a relapse may not always be possible when using a limited number of markers. However, this challenge can be overcome by using polymorphic markers, such as those in the present panel, which offer a theoretical range of 2^7^ to 2^6^ (64-128) possible combinations per locus, based on the median SNP density, providing a high multi-locus resolution. WGS analysis has demonstrated that relapses can be meiotic siblings resulting from the same recombination event, contrary to what microsatellite genotyping classified as identical or clonal.^68^ Recent *P. vivax* wide-genome panels of Ampseq markers could potentially address this question;^6,26,27,29^ however, increasing marker coverage to 10,000 reads would represent higher costs for laboratories in low-resource settings. Using PvAmpSeq data, we estimated IBD and calculated posterior probability estimates of recurrences being relapses, recrudescences or reinfections by using the information from 11 microhaplotype markers. We found that this classification using both methods was largely concordant in samples from both the Solomon Islands clinical trial and the Peruvian community cohort. Nevertheless, simulation studies and/or studies conducted in other epidemiological contexts could inform whether the number of PvAmpSeq markers included in this study would be sufficient for classifying recurrent infections in clinical trials executed in settings with higher endemicity levels of *P. vivax* than in this study.

Despite the advancements offered by high-coverage panels like PvAmpSeq, a major challenge in distinguishing relapses from reinfections remains the inherent limitation of current analytical classification tools (e.g., Pv3Rs). These methods rely on specific assumptions; for example, Pv3Rs assumes recurrence states are either relapse, recrudescence or reinfection where recurrence states are mutually exclusive, assumes no genotyping errors, and does not model persistent infections. For instance, a reinfection may be mistakenly classified as a relapse if the recurrence parasite shows high IBD with the previous infection, as might happen in household transmissions. Conversely, a relapse will have a high reinfection probability if the relapsing parasite exhibits low IBD due to diversity in the hypnozoite reservoir. The Pv3Rs model considers the possibility of residual relapse even if the genetic data are consistent with reinfection (i.e., low IBD), but assumes that recurrence states are mutually exclusive, which may limit applicability in certain settings where recurrence states are liable to overlap. Furthermore, current genotyping approaches are constrained by technical limitations, as they are typically restricted to using DNA derived from blood-stage infections. This approach provides an incomplete picture, as the genetic diversity of the dormant hypnozoite reservoir (the “ground truth” for relapse origin) cannot be directly determined. Given these classification constraints, we adopted a strategy of presenting results from both IBD-based classification methods and model-based classification methods, the integration of which provides a more robust and suitable approach for the analysis of *P. vivax* recurrence patterns.

In conclusion, here we present a new framework to classify *P. vivax* recurrent infections using PvAmpSeq microhaplotype data for molecular correction in drug clinical trials and for studying *P. vivax* relapse biology in longitudinal cohorts, and demonstrate its potential using state-of-the-art analysis methods. We anticipate that this tool can be applied in a wide range of epidemiological studies and clinical trials to fill a pressing need in *P. vivax* genomic epidemiology to better understand relapse epidemiology and support the robust evaluation of clinical efficacy trials.

## Data Availability

All amplicon sequencing data are available under accession no. SAMN43387238 to SAMN43387533 at the Sequence Read Archive (SRA), and the associated BioProject ID is PRJNA1153071. Original data, R scripts, and algorithms developed in this study are accessible in the repository https://github.com/jrosados/PvAmpSeq. De-identified datasets were generated during the current study and used to make all figures available as supplementary files or tables.

https://github.com/jrosados/PvAmpSeq

https://www.ncbi.nlm.nih.gov/bioproject/?term=PRJNA1153071

## Key resources table

| Reagent or Resource | Source | Identifier |
| --- | --- | --- |
| **Bacterial and Virus Strains** |  |  |
| Enterobacteria phage PhiX control v3 | Illumina | Cat# **FC-110-3001** |
| **Chemicals, Peptides, and Recombinant Proteins** |  |  |
| GoTaq® Probe qPCR Master Mix | Promega | Cat# **A6102** |
| Quant-iT PicoGreen® dsDNA kit | Thermo Fisher Scientific | Cat# **P7589** |
| **Critical Commercial Assays** |  |  |
| Favorprep 96-well genomic DNA kit | Favorgen | Cat# **FADWE 96004** |
| E.Z.N.A. Blood DNA Mini Kit | Omega Bio-tek, Inc. | Cat# **D3392-02** |
| QIAGEN MinElute 96 UF PCR Purification Kit | QIAGEN | Cat# **28053** |
| NucleoMag® NGS beads | Macherey-Nagel | Cat# **744970. 50** |
| MiSeq reagent kit v3 (600 cycles; 2 × 300 bp) | Illumina | N/A |
| **Oligonucleotides** |  |  |
| **PvAmpSeq** Primers/Probes (Primary, Secondary, Index) | Eurofins | Sequences provided in https://doi.org/10.5281/zenodo.17474689 |
| Duplex TaqMan qPCR Primers/Probes (Pv and Pf 18s rRNA) | Eurofins | Sequences listed in reference 36 |
| **Software and Algorithms** |  |  |
| R 4.1.0 | The R Foundation | RRID: SCR_001905 |
| AmpSeqR version 0.1 | Bahlolab (via GitHub) | https://github.com/bahlolab/AmpSeqR |
| dcifer R package | Gerlovina I. *et al.* 2022 | https://eppicenter.github.io/dcifer/ |
| Pv3Rs R package | Taylor A.R. *et al.* 2022 | https://github.com/aimeertaylor/Pv3Rs |
| Primer3 | Untergasser, A. *et al.* 2012 | RRID: SCR_003139 |
| Biostrings R package | Pagès, H. *et al.* 2024 | N/A (Standard R package) |
| ShortRead R package | Morgan, M. *et al.* 2009 | N/A (Standard R package) |
| GATK V4 | Broad Institute | RRID: SCR_001876 |
| **Other** |  |  |
| QuantStudio TM 5 Real-Time PCR system | Applied Biosystems | Cat# **A28140** |
| Agilent D5000 ScreenTape System | Agilent Technologies | Cat# **G2991AA** |
| Illumina MiSeq platform | Illumina | N/A |
| PvAmpSeq Genotyping SOP | Zenodo | https://doi.org/10.5281/zenodo.17474689 |

## Acknowledgments

We thank the participants who anonymously agreed to participate in the ACT-Radical clinical trial and the ICEMR study. We acknowledge the field teams that contributed to sample collection in the Solomon Islands and Peru, and Soazic Gardais for logistical support at Institut Pasteur, Paris. We thank Leanne Robinson for provision of the control DNA samples. We thank Prof Miles Markus for his input and suggestions on our manuscript. This work was also made possible through the Victorian State Government Operational Infrastructure Support and Australian Government National Health and Medical Research Council Independent Research Institute Infrastructure Support Scheme. The study was supported by a grant from the Bill and Melinda Gates Foundation (funding grant ID number: OPP1151132). IM was supported by an Australian National Health and Medical Research Council (NHMRC) Investigator Grant (#2016726). MB was supported by an Australian NHMRC Investigator Grant (GNT1195236). The Peruvian cohort was part of the Amazonian International Center of Excellence in Malaria Research that received funding through the Cooperative Agreement U19AI089681 from the US Public Health Service, National Institutes of Health/National Institute of Allergy and Infectious Diseases, USA to JMV, and from a FOGARTY global infectious disease training grant (2D43TW007120-11A1, NIH-USA to JMV). ART is funded by the European Union (project number 101110393). Views and opinions expressed are however those of the author only and do not necessarily reflect those of the European Union or European Research Executive Agency (REA). Neither the European Union nor the granting authority can be held responsible for them. This work was supported by the French government’s “Integrative Biology of Emerging Infectious Diseases” (Investissement d’Avenir grant ANR-10-LABX-62-IBEID) and INCEPTION (Investissement d’Avenir grant ANR-16-CONV-0005) programs to MW. JR postdoctoral fellowship was funded by Institut de Recherche pour le Développement. SR-P acknowledges funding from the MRC Centre for Global Infectious Disease Analysis (reference MR/X020258/1), funded by the UK Medical Research Council (MRC). This UK-funded award is carried out in the frame of the Global Health EDCTP3 Joint Undertaking.

## Author’s contributions

JR, IM, and SRP designed the study. JR and SRP designed PvAmpSeq protocols. JR, ZT, KS, JB and CB did the genotyping. JH, JM, and MB developed AmpSeqR and preprocessed sequencing data, with contributions from JR, SRP, TO and CB. JR, TO, ART and SRP analysed the data and performed statistical analyses. JMV, MTW, MB, DG, and IM provided administrative and logistical support. JR and SRP wrote the first draft of the manuscript. All authors had full access to all the data. All authors contributed to the manuscript’s writing.

## Supplemental information

Document S1. Figures S1–S16

Document S2. Table S1–S9

Text S1. Excel file containing additional data on PvAmpSeq markers, too large to fit in a PDF.

Text S2. Data of IBD estimates in Solomon Islands cohort.

Text S3. Data of IBD estimates in the Peruvian cohort.

