## Supplemental_files for "Understanding *Plasmodium vivax* recurrent infections using an amplicon deep sequencing assay, PvAmpSeq, identity-by-descent and model-based classification": Document S1_REVISED_19022026.pdf

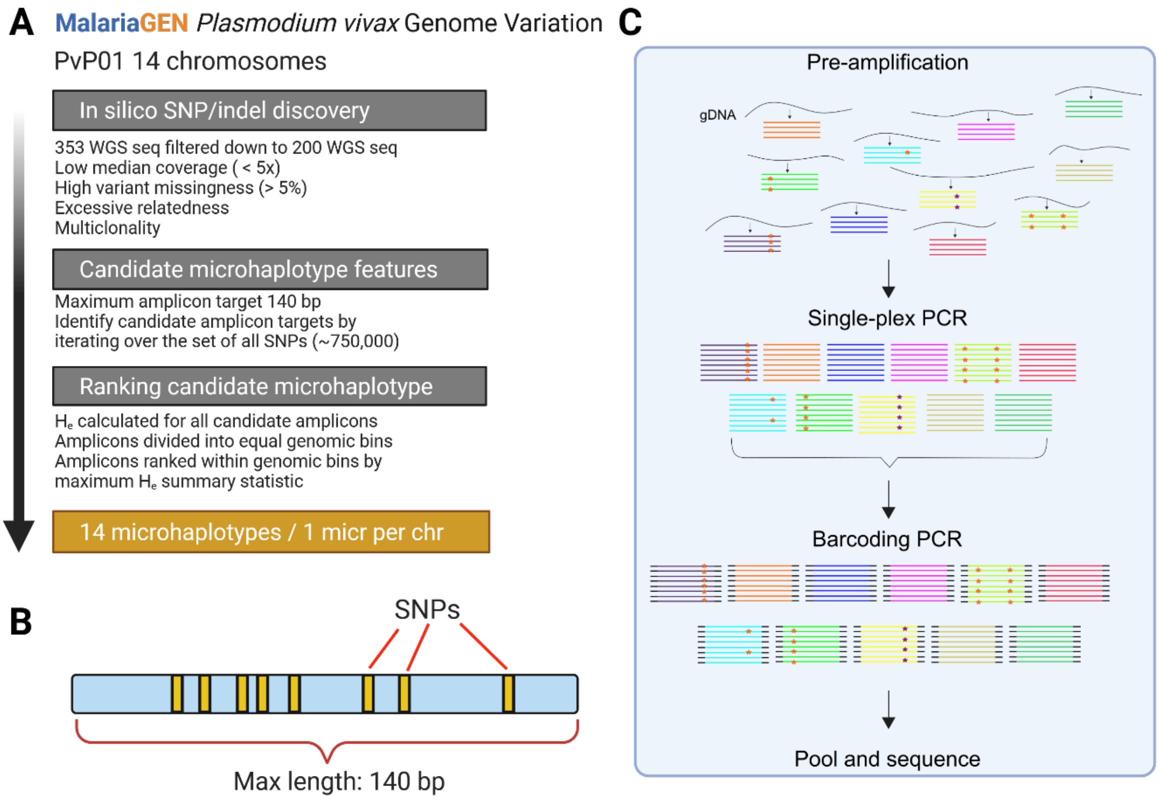


**Figure S1**. Design of PvAmpSeq. (A) Pipeline for the design of the initial 14 microhaplotype markers of PvAmpSeq. WGS: Whole genome sequence. He: Expected heterozygosity. (B) Schematic of a PvAmpSeq microhaplotype displaying SNPs or indels (in yellow) within a window of 140bp. (C) Schematic of PvAmpSeq workflow of 11 AmpSeq markers. Asterisks represent SNPs in the microhaplotype, and every microhaplotype is depicted with a different colour.

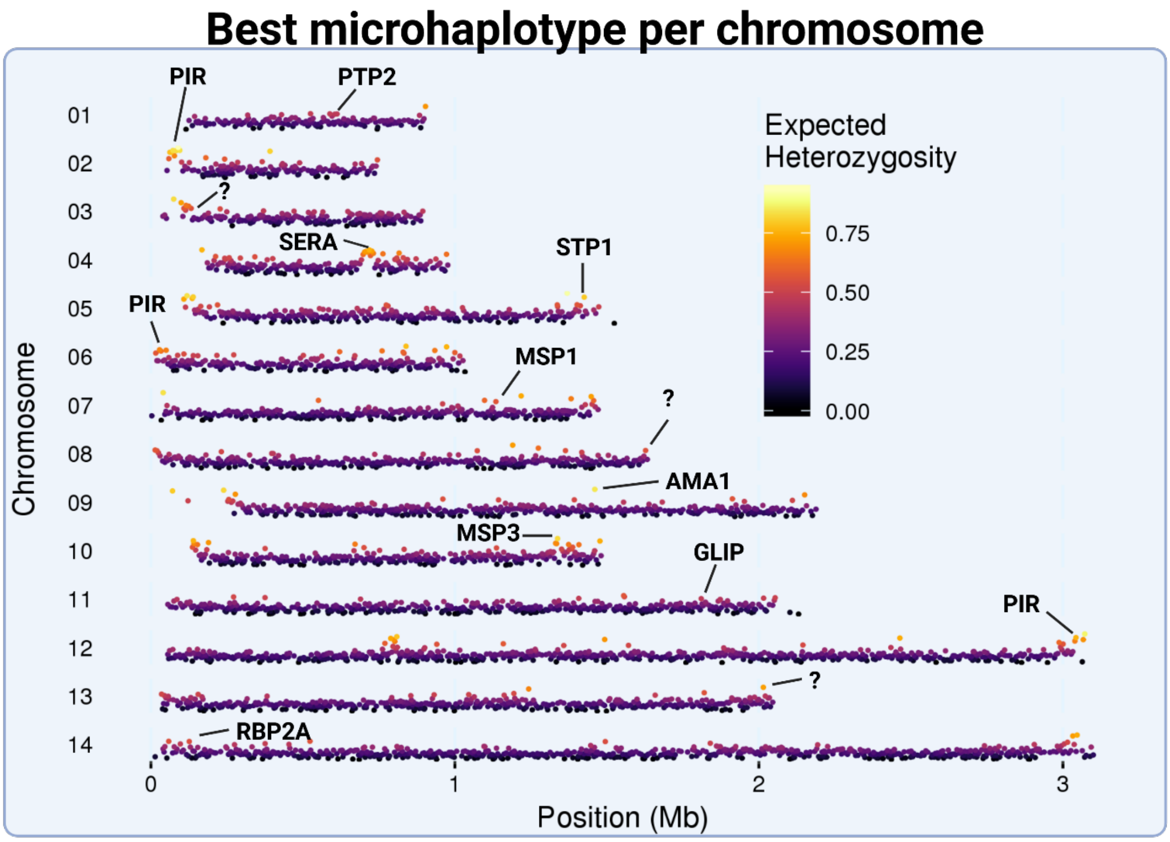


**Figure S2.** Best microhaplotypes per chromosome are depicted on the 14 nuclear chromosomes of the PvP01 reference genome. Microhaplotypes with the highest expected heterozygosity are highlighted with hot colours. PvAmpSeq microhaplotype markers are indicated by black arrows and the name of the gene region. Question marks indicate markers located in gene regions of unknown function.

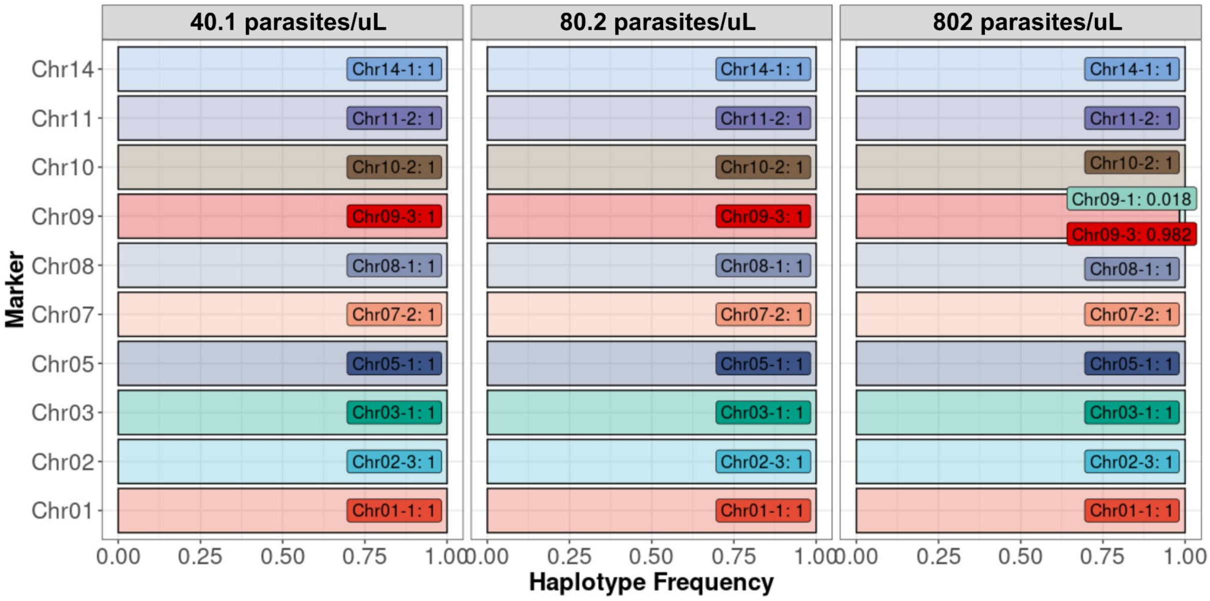


**Figure S3.** Bar plot showing the haplotype frequencies in 10 amplicons of three serial dilutions of *P. vivax* AR-246-0 samples (40.1, 80.2, and 802 parasites/µl). The x-axis represents the haplotype frequency, and the y-axis represents the amplicon marker. Haplotypes and their frequencies are shown in darker-coloured boxes. Haplotype frequencies range from 0.01 to 1.

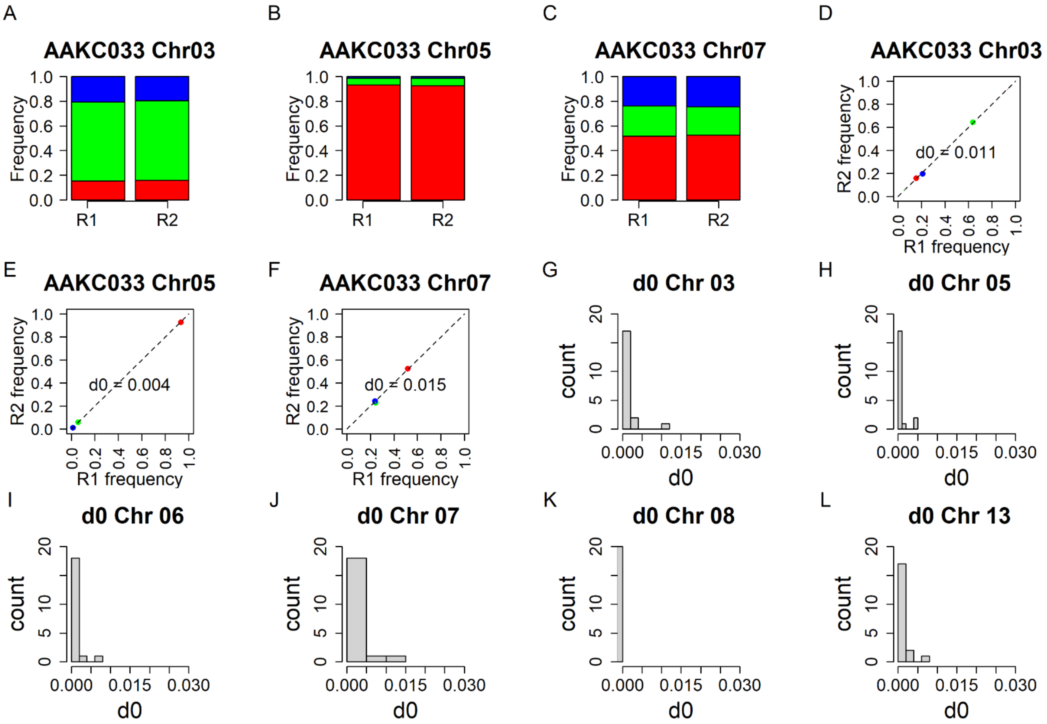


**Figure S4.** Noise and summed distance metric as reported by Mideo *et al*. [1]. A-C) Relative abundance of microhaplotypes detected in technical replicates (R1 and R2) of markers A) Chr03, B) Chr05 and C) Chr07 in patient AAKC033 from Papua New Guinea. D-F) Summed distance metric for microhaplotypes detected in technical replicates of markers D) Chr03, E) Chr05, and F) Chr07 in patient AAKC033. In the barplots and dotplots, each colour represents a unique microhaplotype. Each dot in the dotplot represents the two frequency estimates for each microhaplotype. The dotted lines indicate the distance to the 1:1 line, and the $d_0$ values are the sums of these distances across all microhaplotypes for a given marker within that patient. If technical replicates were perfectly repeatable, all points would lie along the 1:1 line, and *d0* would be 0. G-H) The distribution of *d0* values for Chr03, Chr05, Chr06, Chr07, Chr08 and Chr13 calculated from 20 isolates in the optimisation of PvAmpSeq (Papua New Guinea, n = 8; Peru, n = 12).

1. Mideo N, Kennedy DA, Carlton JM, Bailey JA, Juliano JJ, Read AF. Ahead of the curve: next generation estimators of drug resistance in malaria infections. Trends Parasitol. **2013**; 29(7):321–328.

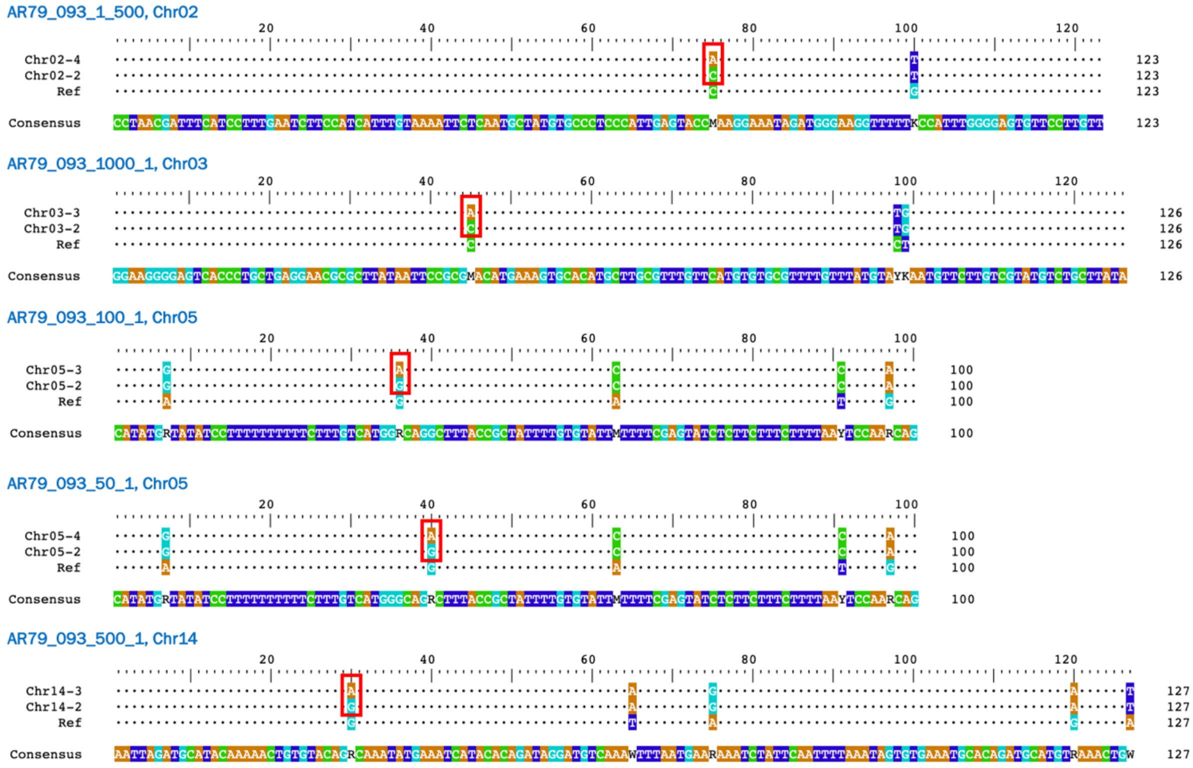


**Figure S5.** Singleton haplotype sequences. Comparison of minority clones and majority clones' haplotype sequences. The name of the haplotypes indicates the name of the marker chromosome and the haplotype number detected in that sample; e.g. Chr14-2. Chr02-4, Chr03-3, Chr05-3, Chr05-4, and Chr14-3 represent minor clones' haplotypes considered singleton after comparing their frequency with the whole dataset. Chr02-2, Chr03-2, Chr05-2, Chr14-2 represent major clones' haplotypes.

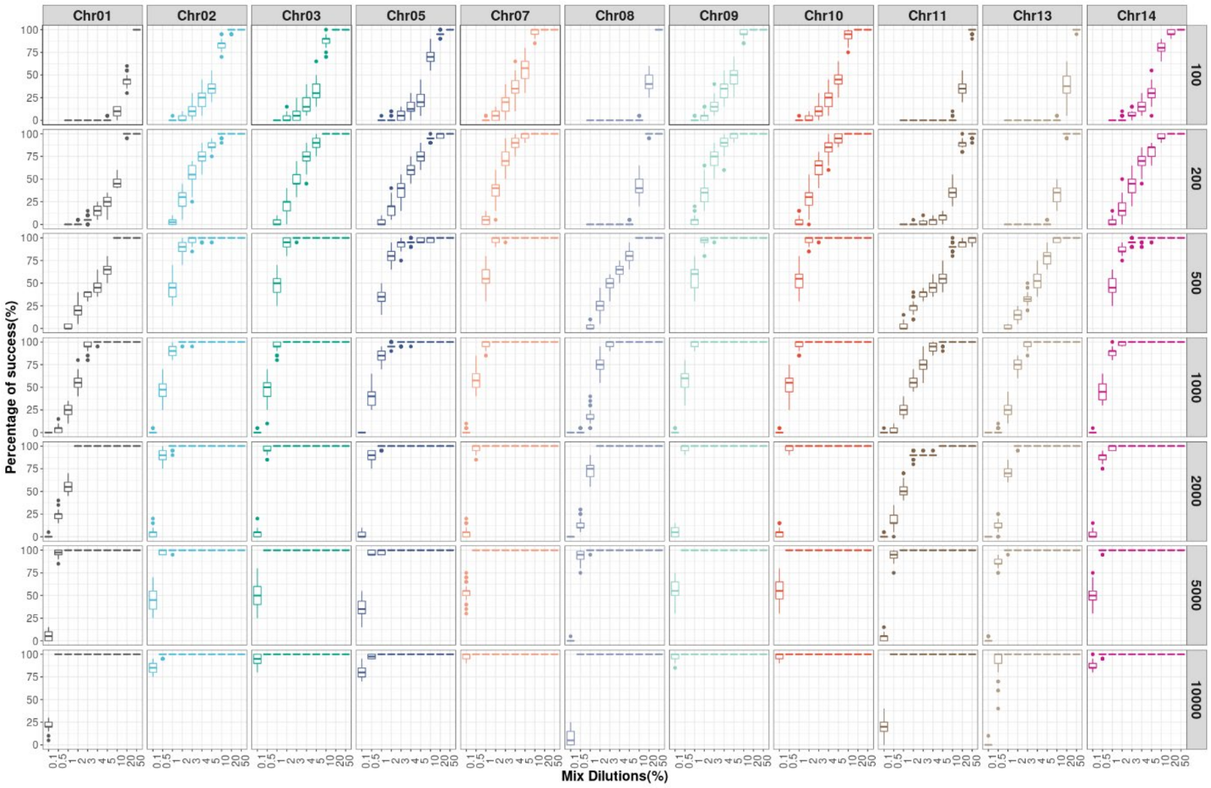


**Figure S6.** The detectability of the minor clone when varying the number of reads and mixture component ratios. The sequences were sampled following the binomial distribution. The x-axis represents the mixture dilutions (%) and the y-axis represents the success rate (%) of detecting the minor clone. Colored by amplicon marker.

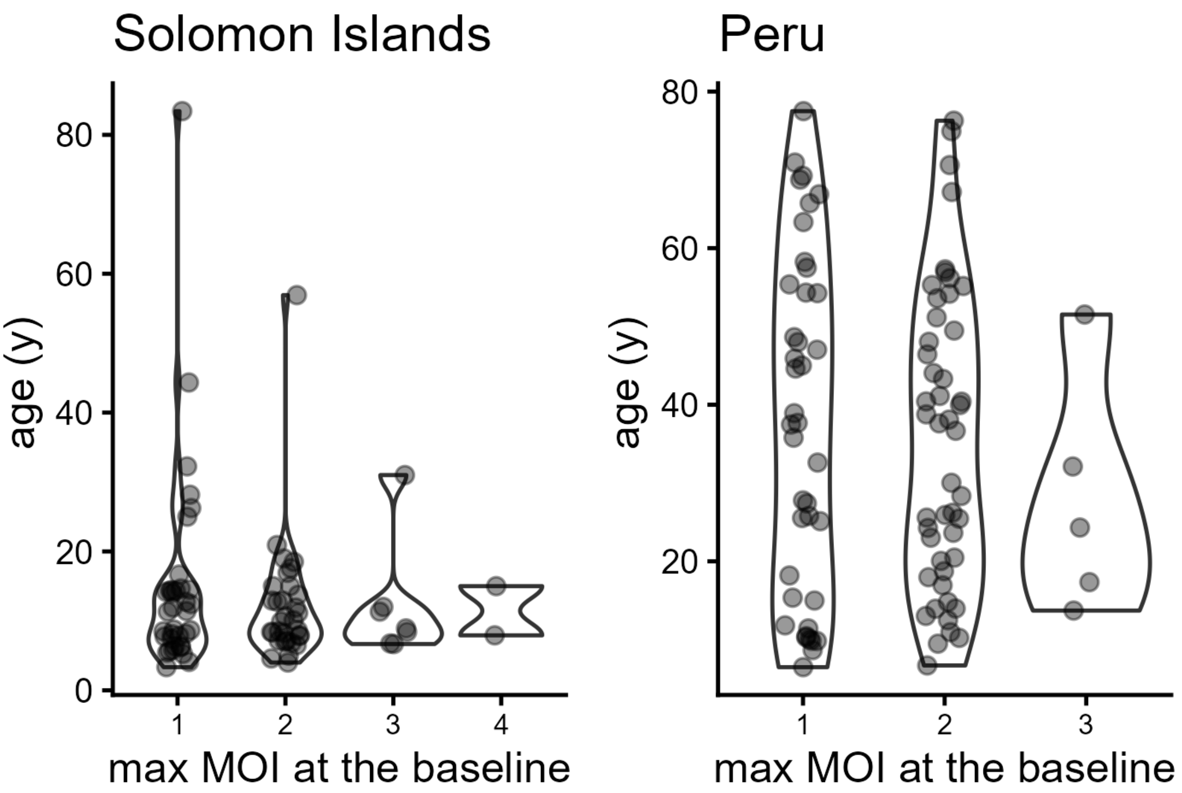


**Figure S7.** Association of age and maximum multiplicity of infection (MOI) in baseline samples. (Kruskal-Wallis test, Solomon Islands $p$ = 0.986, Peru $p$ = 0.672).

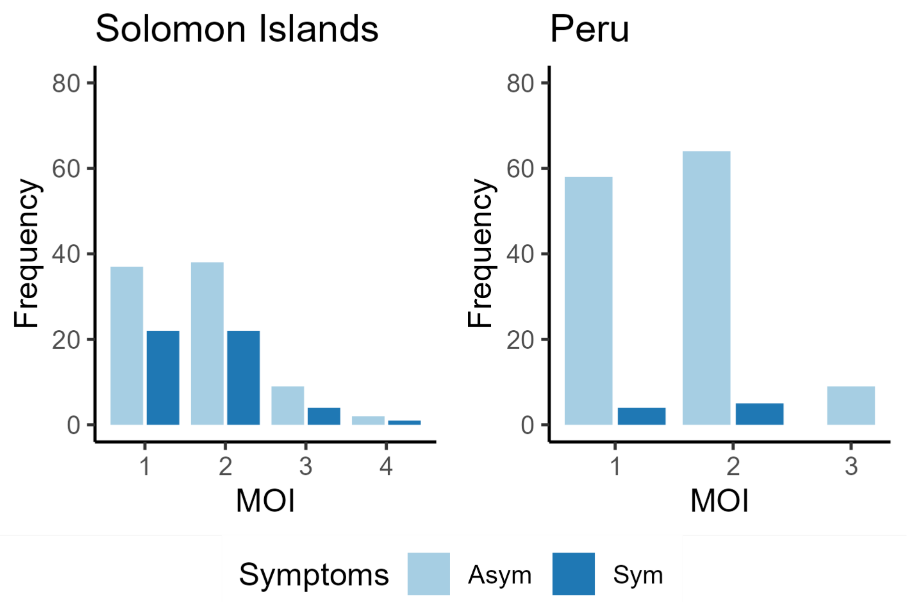


**Figure S8.** Distribution of maximum multiplicity of infection (MOI) detected by all markers in the whole data set of samples sorted by the presence of symptoms. No association was found between the number of clones (MOI) and the presence of symptoms ($\chi 2$ test, Solomon Islands $p = 0.744$, Peru $p = 0.823$)

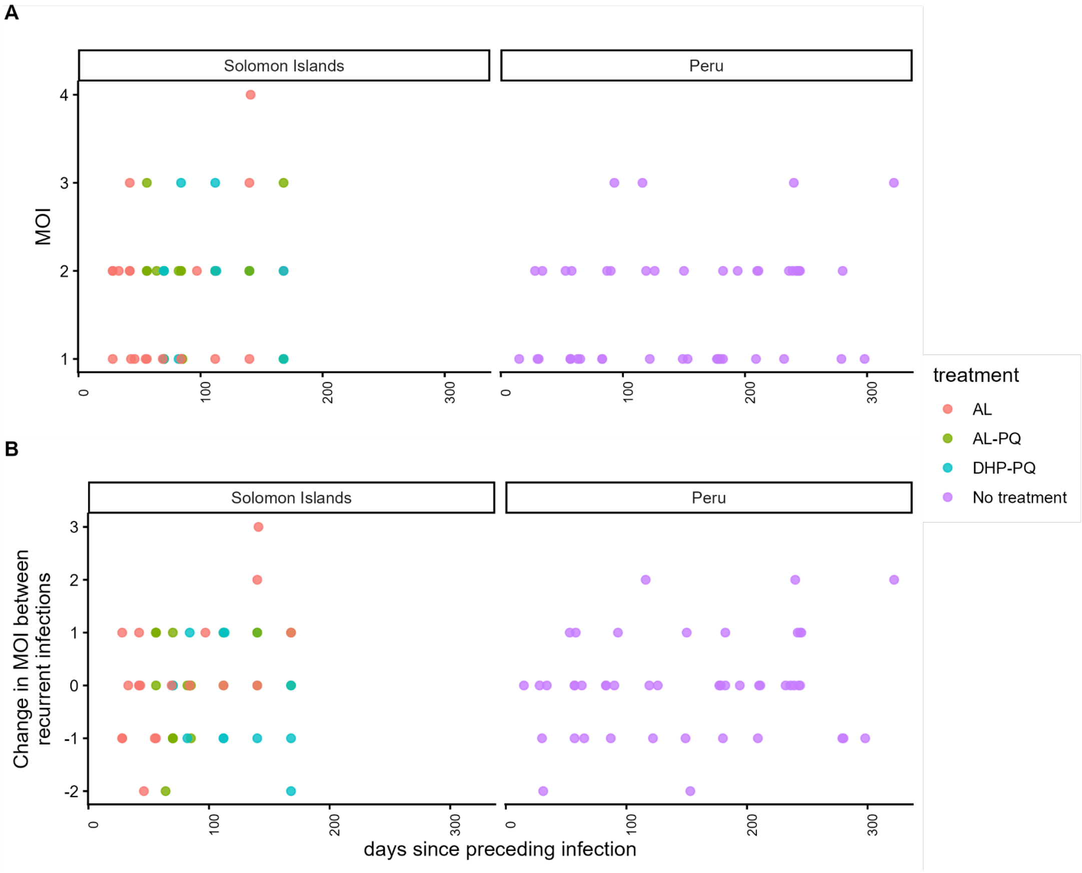


**Figure S9.** Maximum multiplicity of infection (MOI) dynamics over time in Solomon Islands and Peru. (A) Temporal distribution of maximum MOI. Y-axis represents the maximum MOI. X-axis represents the days to next recurrent infection. (B) Change of maximum MOI between recurrent infections over time. Y-axis represents the changes in maximum MOI between recurrent infections. X-axis represents the days since the preceding infection. Infections are represented by dots coloured by treatment type received at day 0. AL: artemether-lumefrantine; AL-PQ: artemether-lumefrantine + primaquine; DHP-PQ : dihydroartemisinin-piperaquine + primaquine.

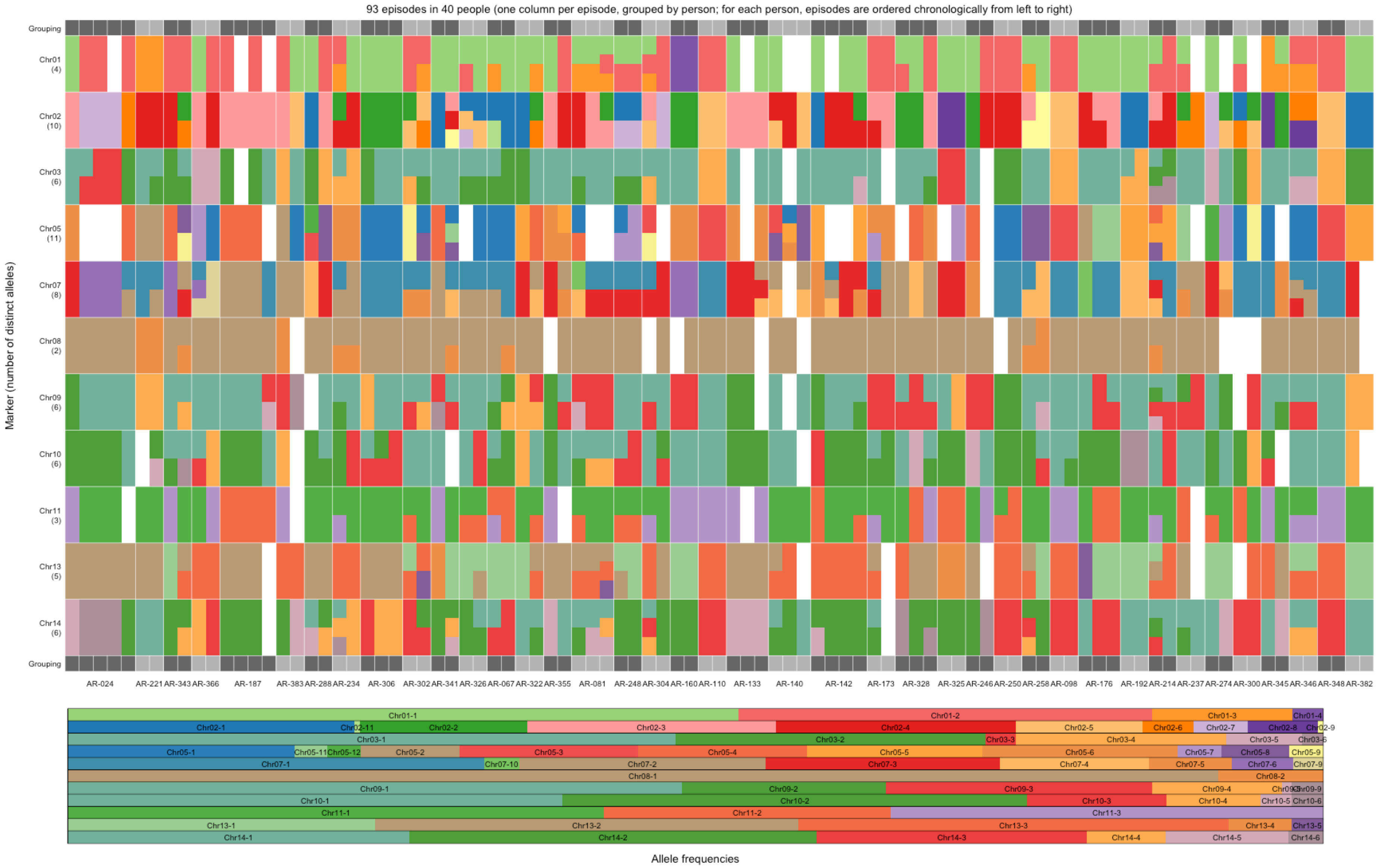


**Figure S10.** Individual-level microhaplotype data and population-level microhaplotype frequencies in the Solomon Islands clinical trial. Upper mosaic plot shows the genotype data of recurrent infections at the individual level. The number of microhaplotypes (alleles) per marker is shown in parentheses. Recurrent episodes are grouped by individual and represented by vertical columns. The bottom horizontal bar plot shows the microhaplotype (allele) frequencies at the population level. Microhaplotypes are coloured according to their relative microhaplotype frequency.

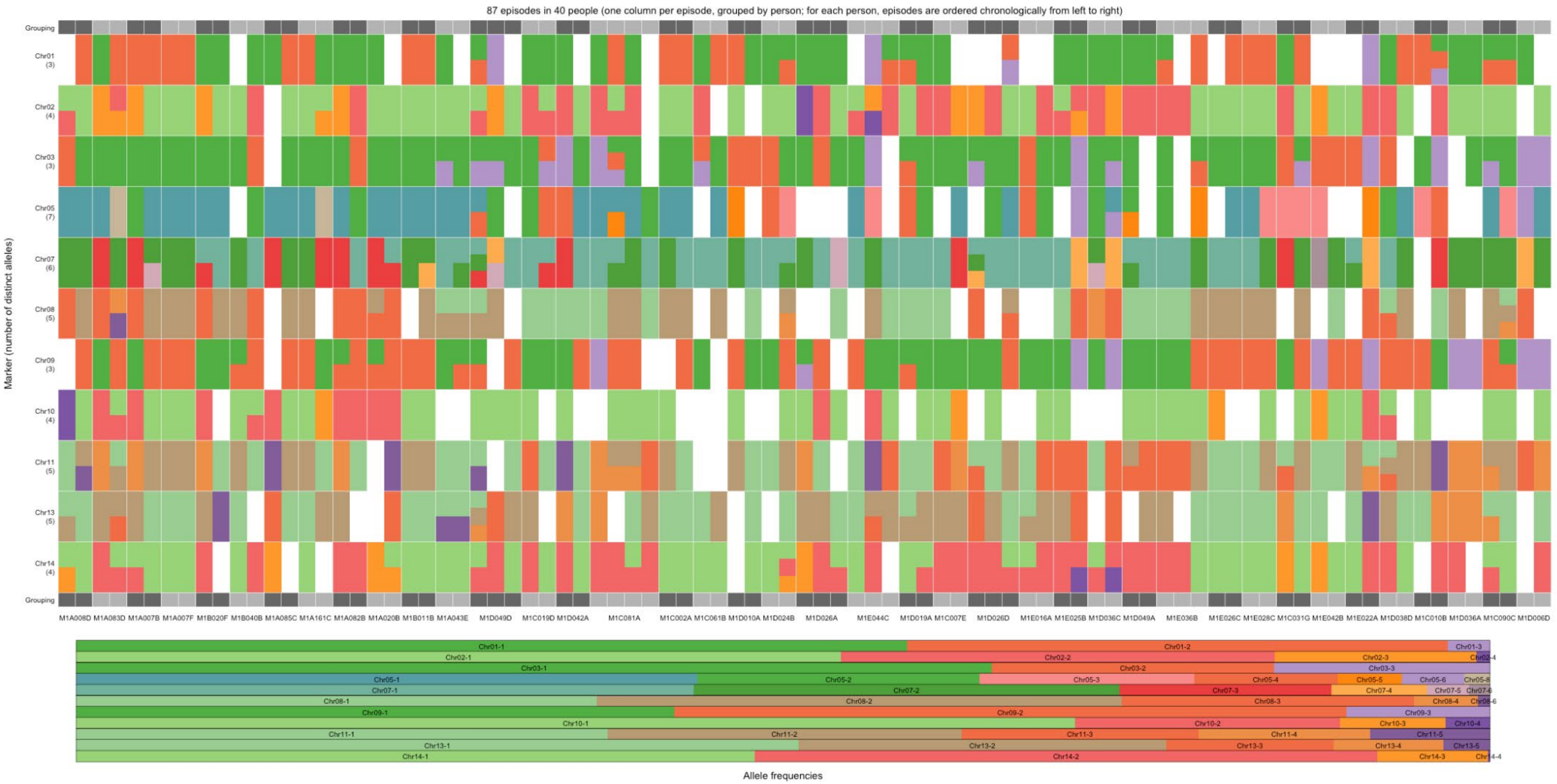


**Figure S11.** Individual-level microhaplotype data and population-level microhaplotype frequencies in the Peru cohort. Upper mosaic plot shows the genotype data of recurrent infections at the individual level. The number of microhaplotypes (alleles) per marker is shown in parentheses. Recurrent episodes are grouped by individual and represented by vertical columns. The bottom horizontal bar plot shows the microhaplotype (allele) frequencies at the population level. Microhaplotypes are coloured according to their relative microhaplotype frequency.

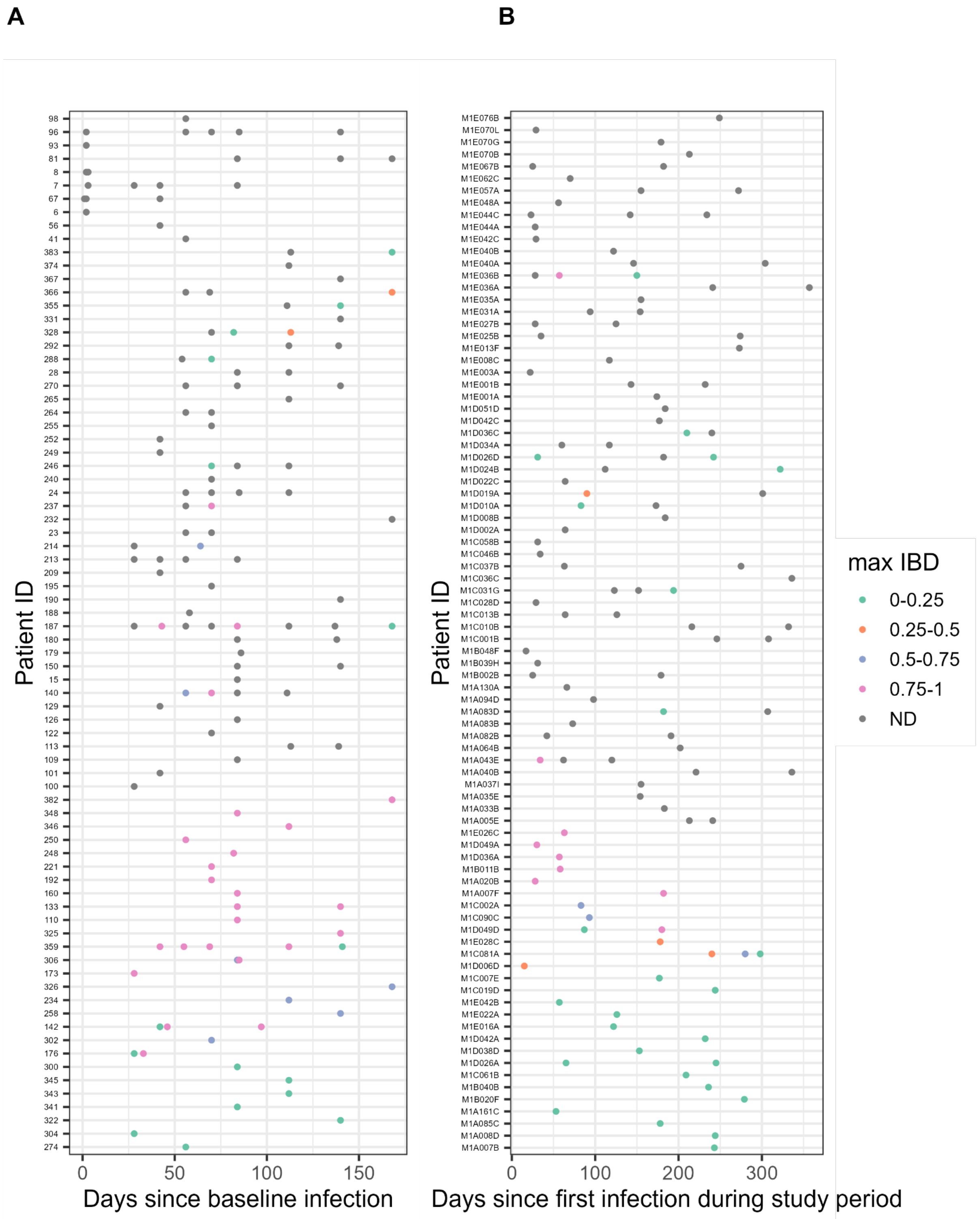


**Figure S12.** Maximum IBD estimate between recurrence and preceding infection pairs in the (A) Solomon Islands and (B) Peru. The X-axis shows the days since baseline (becoming infected), the Y-axis depicts each patient's infection over time, and the colour represents the maximum IBD estimate range for the infection pairs. ND denotes the samples that failed sequencing by PvAmpSeq.

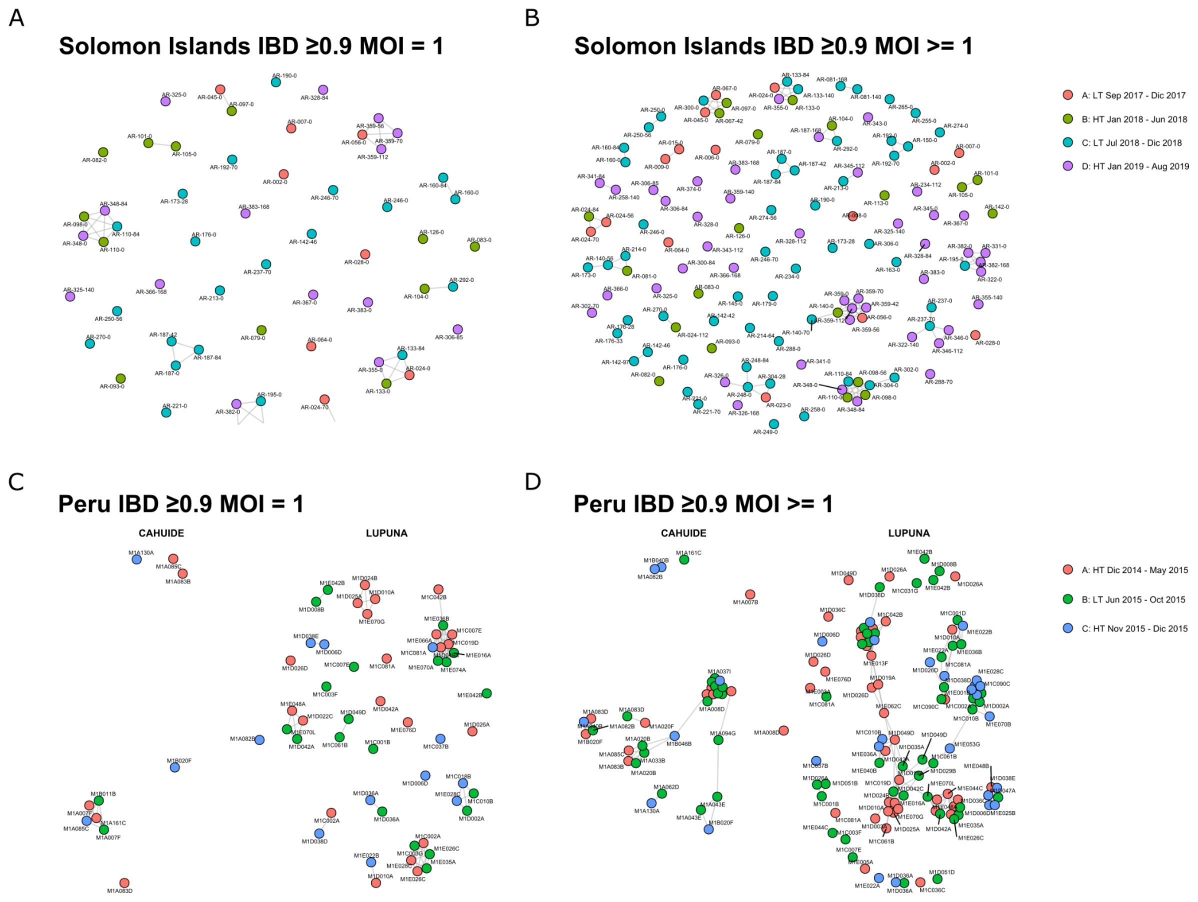


**Figure S13.** Network of *Plasmodium vivax* infections in the Solomon Islands and Peru. Relatedness network for monoclonal samples in (A) the Solomon Islands and (C) Peru at IBD ≥0.9. Relatedness networks for monoclonal and polyclonal samples in the Solomon Islands (B) and Peru (D), respectively, with IBD ≥ 0.9. Nodes (samples) are coloured based on the transmission season of each cohort. Edges show the IBD estimates.

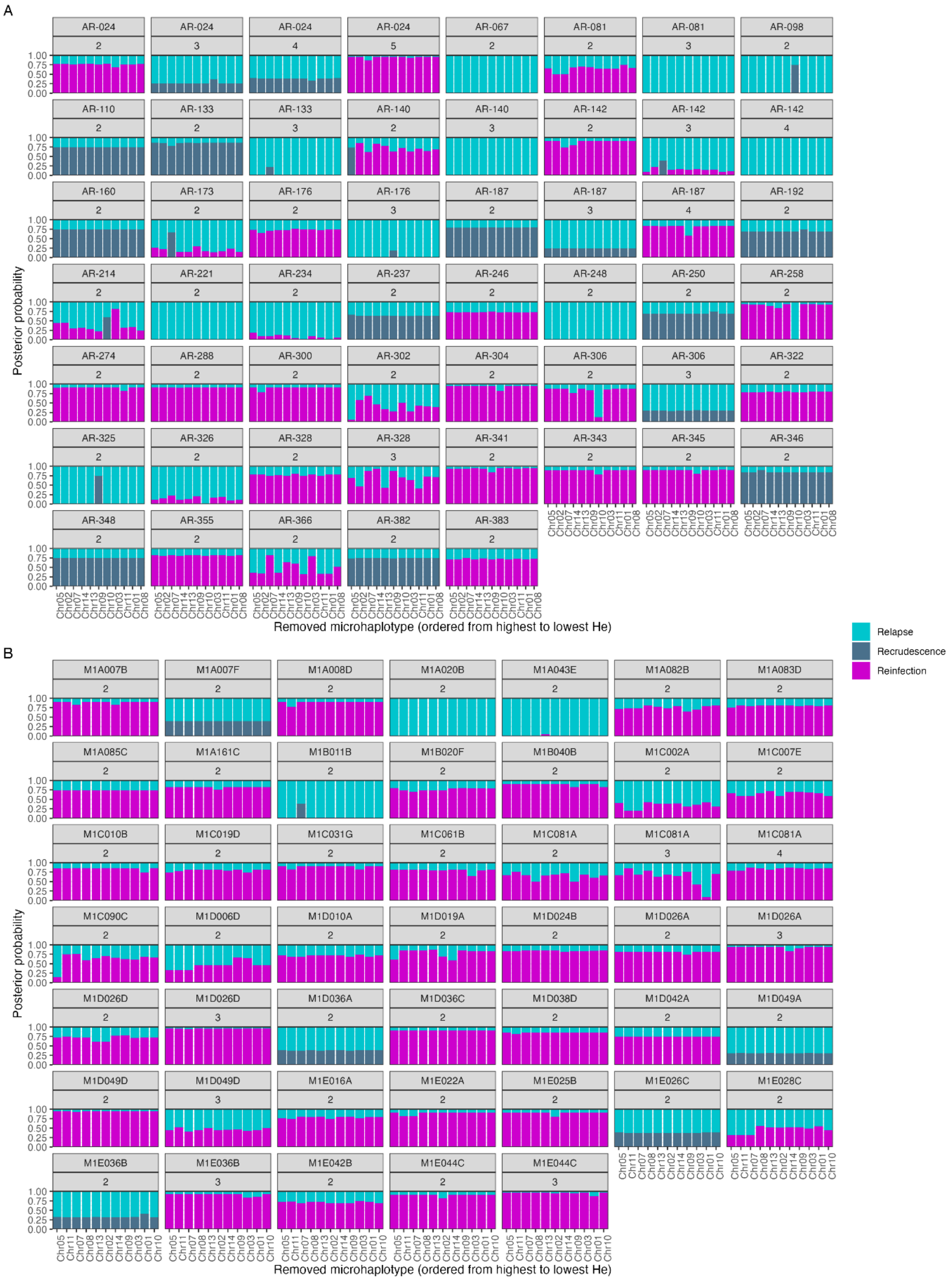


**Figure S14.** Sensitivity analysis of model-based classification of recurrent *P. vivax* infections with PvAmpSeq to assess how particular markers might influence the probabilistic classification results. Posterior probability estimates of recurrence state (relapse, recrudescence or reinfection), faceted by each participant in (A) Solomon Islands and (B) Peru. The X-axis shows the results when a given marker was omitted from the analysis. *Note:* Results that are sensitive to a single omission are likely subject to a genotyping error at that marker.

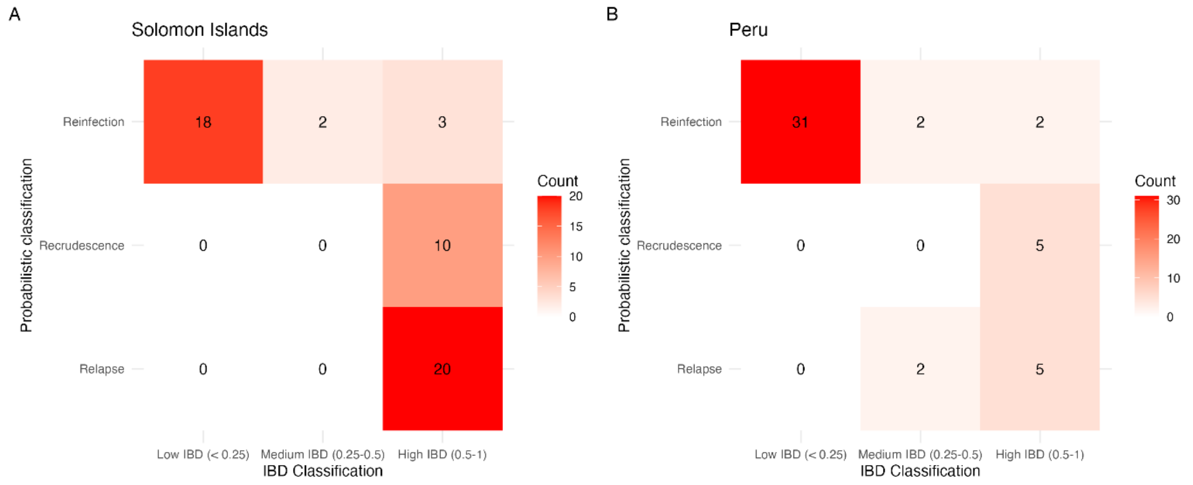


**Figure S15.** Confusion matrix depicting the intersection between identity-by-descent (IBD) classification and the most probable recurrence state given by Pv3Rs in **A)** Solomon Islands and **B)** Peru. IBD metrics were estimated using the *dcifer* R package. IBD ranges: low IBD: 0-0.25; medium IBD: 0.25-0.5; high IBD: 0.5-1. Note that in the case of more than one recurrence, the IBD range refers to the maximum IBD when compared to all preceding infections. For the Peru data, due to a Pv3Rs model misspecification, recurrence state definitions are re-interpreted as follows recrudescence = persistence without added relapse or reinfection; reinfection = reinfection without persistence; relapse = everything else, e.g. (relapse without persistence) OR (persistence AND reinfection) etc (see Methods).

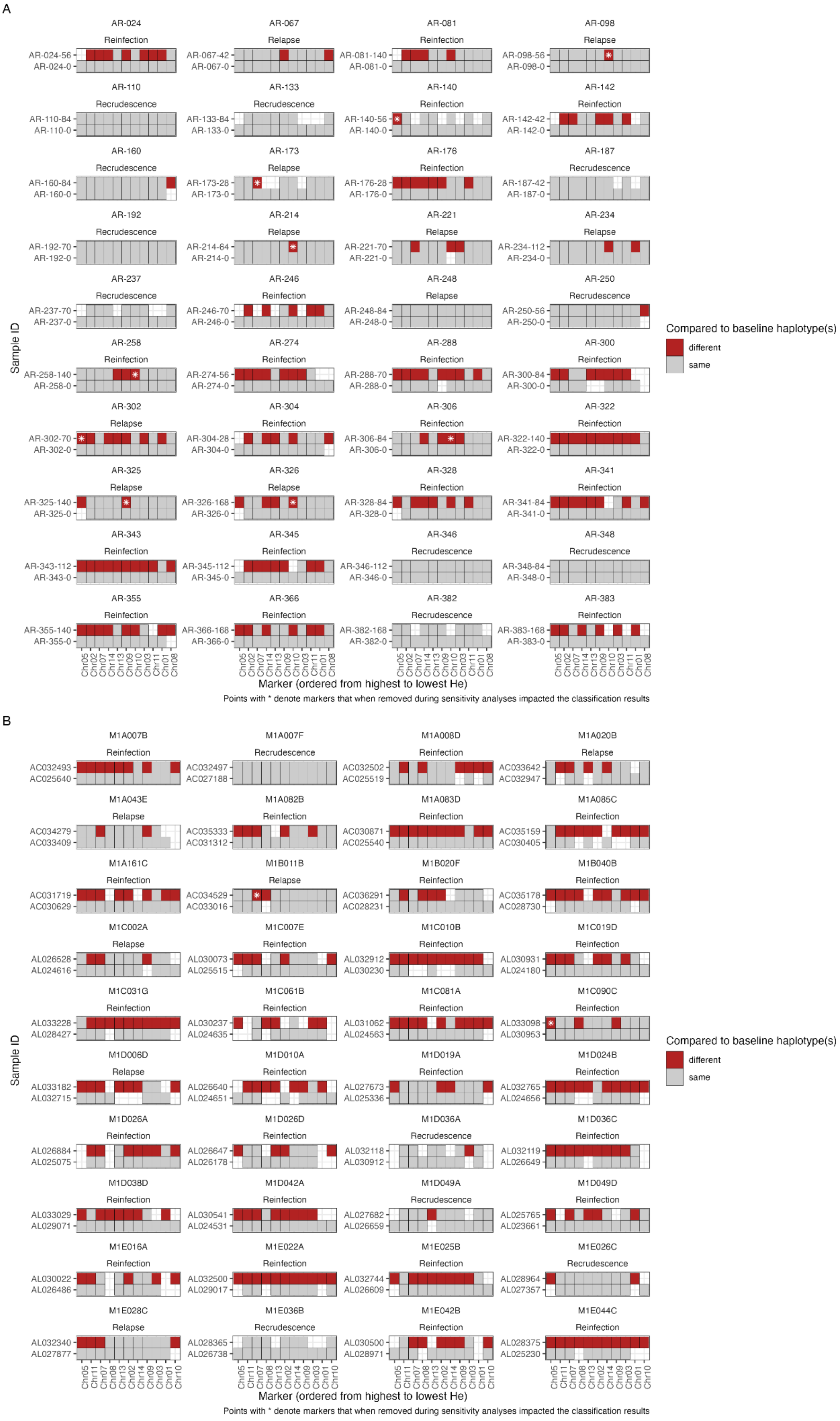
A
AR-024 Reinfection AR-024-56 AR-024-0
AR-067 Relapse AR-067-42 AR-067-0
AR-081 Reinfection AR-081-140 AR-081-0
AR-098 Relapse AR-098-56 AR-098-0
AR-110 Recrudescence AR-110-84 AR-110-0
AR-133 Recrudescence AR-133-84 AR-133-0
AR-140 Reinfection AR-140-56 AR-140-0
AR-142 Reinfection AR-142-42 AR-142-0
AR-160 Recrudescence AR-160-84 AR-160-0
AR-173 Relapse AR-173-28 AR-173-0
AR-176 Reinfection AR-176-28 AR-176-0
AR-187 Recrudescence AR-187-42 AR-187-0
AR-192 Recrudescence AR-192-70 AR-192-0
AR-214 Relapse AR-214-64 AR-214-0
AR-221 Relapse AR-221-70 AR-221-0
AR-234 Relapse AR-234-112 AR-234-0
AR-237 Recrudescence AR-237-70 AR-237-0
AR-246 Reinfection AR-246-70 AR-246-0
AR-248 Relapse AR-248-84 AR-248-0
AR-250 Recrudescence AR-250-56 AR-250-0
AR-258 Reinfection AR-258-140 AR-258-0
AR-274 Reinfection AR-274-56 AR-274-0
AR-288 Reinfection AR-288-70 AR-288-0
AR-300 Reinfection AR-300-84 AR-300-0
AR-302 Relapse AR-302-70 AR-302-0
AR-304 Reinfection AR-304-28 AR-304-0
AR-306 Reinfection AR-306-84 AR-306-0
AR-322 Reinfection AR-322-140 AR-322-0
AR-325 Relapse AR-325-140 AR-325-0
AR-326 Relapse AR-326-168 AR-326-0
AR-328 Reinfection AR-328-84 AR-328-0
AR-341 Reinfection AR-341-84 AR-341-0
AR-343 Reinfection AR-343-112 AR-343-0
AR-345 Reinfection AR-345-112 AR-345-0
AR-346 Recrudescence AR-346-112 AR-346-0
AR-348 Recrudescence AR-348-84 AR-348-0
AR-355 Reinfection AR-355-140 AR-355-0
AR-366 Reinfection AR-366-168 AR-366-0
AR-382 Recrudescence AR-382-168 AR-382-0
AR-383 Reinfection AR-383-168 AR-383-0
Sample ID
Chr05 Chr02 Chr07 Chr14 Chr13 Chr09 Chr10 Chr03 Chr11 Chr01 Chr08
Marker (ordered from highest to lowest He)
Points with * denote markers that when removed during sensitivity analyses impacted the classification results
Compared to baseline haplotype(s)
different
same
B
M1A007B Reinfection AC032493 AC025640
M1A007F Recrudescence AC032497 AC027188
M1A008D Reinfection AC032502 AC025519
M1A020B Relapse AC033642 AC032947
M1A043E Relapse AC034279 AC033409
M1A082B Reinfection AC035333 AC031312
M1A083D Reinfection AC030871 AC025540
M1A085C Reinfection AC035159 AC030405
M1A161C Reinfection AC031719 AC030629
M1B011B Relapse AC034529 AC033016
M1B020F Reinfection AC036291 AC028231
M1B040B Reinfection AC035178 AC028730
M1C002A Relapse AL026528 AL024616
M1C007E Reinfection AL030073 AL025515
M1C010B Reinfection AL032912 AL030230
M1C019D Reinfection AL030931 AL024180
M1C031G Reinfection AL033228 AL028427
M1C061B Reinfection AL030237 AL024635
M1C081A Reinfection AL031062 AL024563
M1C090C Reinfection AL033098 AL030953
M1D006D Relapse AL033182 AL032715
M1D010A Reinfection AL026640 AL024651
M1D019A Reinfection AL027673 AL025336
M1D024B Reinfection AL032765 AL024656
M1D026A Reinfection AL026884 AL025075
M1D026D Reinfection AL026647 AL026178
M1D036A Recrudescence AL032118 AL030912
M1D036C Reinfection AL032119 AL026649
M1D038D Reinfection AL033029 AL029071
M1D042A Reinfection AL030541 AL024531
M1D049A Recrudescence AL027682 AL026659
M1D049D Reinfection AL025765 AL023661
M1E016A Reinfection AL030022 AL026486
M1E022A Reinfection AL032500 AL029017
M1E025B Reinfection AL032744 AL026609
M1E026C Recrudescence AL028964 AL027357
M1E028C Relapse AL032340 AL027877
M1E036B Recrudescence AL028365 AL026738
M1E042B Reinfection AL030500 AL028971
M1E044C Reinfection AL028375 AL025230
Sample ID
Chr05 Chr11 Chr07 Chr08 Chr13 Chr02 Chr14 Chr09 Chr03 Chr01 Chr10
Marker (ordered from highest to lowest He)
Points with * denote markers that when removed during sensitivity analyses impacted the classification results
Compared to baseline haplotype(s)
different
same

**Figure S16.** Microhaplotype data comparing recurrent infection(s) to the baseline in (A) Solomon Islands and (B) Peru. Samples are grouped by patient and annotated by the model-based recurrence classification. For each patient, the bottom bar shows the baseline episode and the top bar(s) the first recurrent episode. The x-axis shows markers ordered by decreasing expected heterozygosity (He). Grey boxes indicate microhaplotypes shared with the baseline infection, and red boxes denote microhaplotypes that differ from the baseline. Note: this simplifies the overall trends, particularly when the sample is MOI>1, because any haplotype that differs from baseline is coloured red, even if multiple haplotypes are possible (including the same as baseline). Asterisks (*) show markers whose removal during sensitivity analyses changed the recurrence classification (see Figure S12).
