## Supplemental_files for "Understanding *Plasmodium vivax* recurrent infections using an amplicon deep sequencing assay, PvAmpSeq, identity-by-descent and model-based classification": Document S2_REVISED_19022026.pdf

**Table S1.** Comparison of existing *Plasmodium vivax* amplicon sequencing panels

| Method | Key Use Case | Amplicons (or MIPs) | Neutral SNPs | Drug resistance | Vaccine candidates | Advantages | Disadvantages | Type of sample | LOD | Minority clones LOD | Library preparation | Enrichment method* | Depth per marker¶ | Marker success rate | Samples | Samples origin |
|---|---|---|---|---|---|---|---|---|---|---|---|---|---|---|---|---|
| Pv Ampliseq | Population structure, transmission networks, geographical origin of imported cases, drug resistance | 229 | No | 11 genes | 1 gene | Multiplexed, high resolution within high transmission setting, alllows IBD estimation, clonality | Proprietary library kit, requires updating for new geographical contexts | DBS | 5p/uL | 10% | AmpliSeq Library PLUS (Illumina) | sWGA | ≥100 | 95% | 624 | Vietnam, Peru |
| MIP | Population structure, drug resistance, vaccine targets, CNV, signatures of selection | 645 (PvG), 794 (FST), 306 (MAF5), 426 (MAF40) | Yes | 4 genes | 13 genes | Multiplexed, broad geographical applicability, high resolution within high transmission setting, alllows IBD estimation, polyclonality | High limit of detection | DBS | 100p/uL | NA | non-propietary | no | >3 UMI | 69-85% | 685-697 | Peru |
| Kleinecke | Population genetics, transmission networks, distinguish relapse and recrudescence from reinfections in TES | 98 | Yes | 4 genes | No | Multiplexed, high resolution within high transmission setting, alllows IBD estimation, polyclonality | Proprietary library kit | WBS/DBS | 70p/uL | NA | rhAmpSeq library kit(IDT) | no | ≥25 | >86% | 705 | Afghanistan, Bangladesh, Colombia, Ethiopia, Indonesia, and Vietnam |
| PvGAP | Population genetics, transmission networks, geographical origin of imported cases, drug resistance | 88 | Yes | 8 genes | No | Multiplexed, broad geographical applicability, high resolution within high transmission setting, alllows IBD estimation, polyclonality | High limit of detection | WBS/DBS | 100p/ul | NA | non-proprietary | sWGA | ≥10 | 75% | 48 | Ethiopia |
| PvAmpSeq | Distinguish relapse and recrudescence from reinfections in TES, transmission networks | 11 | No | No | No | High resolution within high transmission setting, alllows IBD estimation, polyclonality, designed for TES | Needs to be updated to a multiplexed format | WBS/DBS | 10p/uL | 2% | non-proprietary | Pre-Amp | ≥1000 | 92% | 275 | Solomon Islands, Papua New Guinea, Peru |

TES: Therapeutic efficacy studies; DBS: Dried blood samples; WBS: Whole blood samples; LOD: Limit of Detection; *: Enrichment method for low parasitaemia samples; ¶: Recommended number of reads per marker or Unique Molecular Identifier (UMI).

**Table S2.** Success rate of markers after excluding PCR artefacts

| Marker | Solomon Islands | | | Peru | | |
|---|---|---|---|---|---|---|
| | **Success rate baseline (n = 77)** | **Success rate follow-up (n = 58)** | **Success rate all samples (n = 135)** | **Success rate baseline (n = 93)** | **Success rate follow-up (n = 47)** | **Success rate all samples (n = 140)** |
| Chr01-PTP2 | 0.99 | 0.83 | 0.92 | 0.78 | 0.77 | 0.77 |
| Chr02-Lysophospholipase, putative | 1.00 | 1.00 | 1.00 | 0.98 | 0.94 | 0.96 |
| Chr03-conserved Plasmodium protein | 1.00 | 0.91 | 0.96 | 0.99 | 0.89 | 0.96 |
| Chr05-STP1 | 0.88 | 0.86 | 0.87 | 0.82 | 0.81 | 0.82 |
| Chr07-MSP1 | 1.00 | 0.95 | 0.98 | 1.00 | 1.00 | 1.00 |
| Chr08-Plasmodium exported protein | 0.90 | 0.90 | 0.90 | 0.76 | 0.83 | 0.79 |
| Chr09-AMA1 | 0.99 | 0.95 | 0.96 | 0.99 | 0.91 | 0.96 |
| Chr10-MSP3.3 | 0.96 | 0.91 | 0.94 | 0.70 | 0.70 | 0.70 |
| Chr11-GLIP | 1.00 | 0.93 | 0.96 | 0.98 | 0.94 | 0.96 |
| Chr13-conserved Plasmodium protein | 0.99 | 0.91 | 0.96 | 0.93 | 0.87 | 0.91 |
| Chr14-RBP2a | 1.00 | 0.97 | 0.99 | 0.98 | 0.89 | 0.95 |

The success rate was estimated by dividing the number of samples that remained after quality control filtering by the total number of expected samples.

**Table S3.** Genetic complexity of samples estimated by individual markers

| Marker | Solomon Islands | | | | | | Peru | | | | | |
|---|---|---|---|---|---|---|---|---|---|---|---|---|
| | Baseline samples (n = 77) | | | Follow-up samples (n = 58) | | | Baseline samples (n = 93) | | | Follow-up samples (n = 47) | | |
| | mean MOI (min-max) | Proportion of polyclonal (%) | Number of alleles | mean MOI (min-max) | Proportion of polyclonal (%) | Number of alleles | mean MOI (min-max) | Proportion of polyclonal (%) | Number of alleles | mean MOI (min-max) | Proportion of polyclonal (%) | Number of alleles |
| Chr01-PTP2 | 1.18 (1-3) | 13/76 (17.10) | 2 | 1.19 (1-3) | 8/48 (16.66) | 2 | 1.14 (1-2) | 10/69 (14.49) | 3 | 1.17 (1-3) | 5/36 (13.88) | 3 |
| Chr02-Lysophospholipase, putative | 1.26 (1-3) | 19/77 (24.67) | 13 | 1.26 (1-3) | 13/59 (22.41) | 13 | 1.17 (1-2) | 15/87 (17.24) | 10 | 1.14 (1-2) | 6/44 (13.63) | 10 |
| Chr03-conserved Plasmodium protein | 1.22 (1-3) | 16/77 (20.77) | 8 | 1.19 (1-2) | 10/53 (18.87) | 8 | 1.16 (1-3) | 13/88 (14.77) | 4 | 1.17 (1-3) | 6/42 (14.28) | 4 |
| Chr05-STP1 | 1.18 (1-3) | 11/68 (16.17) | 7 | 1.16 (1-3) | 6/50 (12.00) | 7 | 1.12 (1-2) | 9/73 (12.33) | 6 | 1.05 (1-2) | 2/38 (5.26) | 6 |
| Chr07-MSP1 | 1.27 (1-3) | 18/77 (23.37) | 43 | 1.38 (1-3) | 18/55 (32.73) | 43 | 1.20 (1-3) | 14/89 (15.73) | 34 | 1.17 (1-2) | 8/47 (17.02) | 34 |
| Chr08-Plasmodium exported protein | 1.07 (1-2) | 5/69 (7.24) | 1 | 1.10 (1-2) | 5/52 (9.61) | 1 | 1.16 (1-2) | 11/68 (16.17) | 13 | 1.15 (1-3) | 5/39 (12.80) | 13 |
| Chr09-AMA1 | 1.27 (1-4) | 17/75 (22.66) | 12 | 1.31 (1-2) | 17/55 (30.91) | 13 | 1.14 (1-2) | 12/89 (13.63) | 9 | 1.05 (1-2) | 2/43 (4.65) | 9 |
| Chr10-MSP3.3 | 1.24 (1-4) | 16/74 (21.62) | 14 | 1.21 (1-2) | 11/53 (20.75) | 14 | 1.13 (1-2) | 8/62 (12.90) | 11 | 1.06 (1-2) | 2/33 (6.06) | 12 |
| Chr11-GLIP | 1.18 (1-2) | 14/77 (18.18) | 2 | 1.11 (1-2) | 6/53 (11.32) | 2 | 1.19 (1-3) | 16/87 (18.40) | 5 | 1.11 (1-2) | 5/44 (11.36) | 5 |
| Chr13-conserved Plasmodium protein | 1.14 (1-3) | 9/76 (11.84) | 4 | 1.17 (1-3) | 8/53 (15.09) | 4 | 1.16 (1-3) | 11/83 (13.25) | 3 | 1.07 (1-2) | 3/41 (7.32) | 3 |
| Chr14-RBP2a | 1.26 (1-3) | 16/77 (20.77) | 5 | 1.27 (1-4) | 11/56 (19.64) | 5 | 1.18 (1-2) | 16/87 (18.39) | 4 | 1.24 (1-3) | 9/42 (22.43) | 4 |

Samples with more than one parasite clone were defined as polyclonal samples. Alleles per marker included SNPs, INS and DEL. MOI: Multiplicity of infection or number of parasite clones.

**Table S4.** Multiplicity of infections estimated by combined markers in paired infections

| | Cohort | Baseline samples | Follow-up samples | p-value |
|---|---|---|---|---|
| **mean MOI (min-max)** [a] | Solomon Islands | 1.83 (1-4) | 1.74 (1-4) | p>0.05 |
| | Peru | 1.62 (1-3) | 1.62 (1-3) | p>0.05 |
| **Polyclonal infections (%)** [b] | Solomon Islands | 26/41 (63.4%) | 35/58 (60.3%) | p>0.05 |
| | Peru | 22/40 (55.0%) | 25/47 (53.2%) | p>0.05 |

MOI: Multiplicity of infection or the number of parasite clones (haplotypes).

Samples with more than one parasite clone were defined as polyclonal samples.

a: Two-sided Mann-Whitney test

b: $\chi 2$ test

**Table S5.** Comparison of AmpSeq genotyping versus Microsatellite genotyping in Peruvian samples

| Samples (n = 5) | AmpSeq | | | Microsatellites | | |
|---|---|---|---|---|---|---|
| | MOI | Polymorphic markers | Number markers | MOI | Polymorphic markers | Number markers |
| AL023630 | 1 | 0 | 10 | Monoclonal | 0 | 16 |
| AL023661 | 3 | 10 | 11 | Polyclonal | 4 | 16 |
| AL025728 | 2 | 3 | 11 | Monoclonal | 0 | 16 |
| AL025736 | 1 | 0 | 8 | Monoclonal | 0 | 9 |
| AL025765 | 2 | 2 | 9 | Monoclonal | 0 | 16 |

MOI: Multiplicity of infection. Polymorphic markers: Number of markers that detected more than one clone. Number markers: Number of markers successfully genotyped. AmpSeq genotype data were generated in this study. Microsatellite genotype data were retrieved from Manrique et al. 2019. PLoS Negl Trop Dis. doi: 10.1371/journal.pntd.0007876

**Table S6.** Association between clonality, treatment and IBD of recurrent infections

| | Solomon Islands | | | | Peru | | | |
|---|---|---|---|---|---|---|---|---|
| | **IBD ≤0.25** | **IBD 0.25—0.5** | **IBD ≥0.5** | **p-value** | **IBD ≤0.25l** | **IBD 0.25—0.5** | **IBD ≥0.5** | **p-value** |
| **Clonality** | | | | | | | | |
| Monoclonal, number | 6 | 1 | 16 | $p > 0.05$ [a] | 13 | 2 | 7 | $p > 0.05$ |
| Polyclonal, number | 18 | 1 | 16 | | 20 | 2 | 4 | |
| **Treatment** | | | | | | | | |
| AL, number | 7 | 0 | 15 | $p > 0.05$ [b] | | | | |
| PQ, number | 17 | 2 | 17 | | | | | |

a: Fisher exact test was used to compare the frequency of polyclonal and monoclonal infections in each IBD range group.

b: Fisher exact test was used to compare the frequency of high and low IBD by treatment scheme. AL: artemether-lumefrantine only (n=22); PQ: Included participants who received artemether-lumefrantine + primaquine (AL-PQ, n=22) and participants who received dihydroartemisinin-piperaquine + primaquine (DHP-PQ, n=14).

**Table S7.** Prior probability distribution in Solomon Islands

| **Treatment arm** | **Days since baseline treatment** | **Prior probability of recrudescence (C)** | **Prior probability of relapse (L)** | **Prior probability of reinfection (I)** |
|---|---|---|---|---|
| AL | ≤30 | 0.05 | 0.65 | 0.3 |
| PQ *(AL+PQ and DP+PQ)* | ≤30 | 0.05 | 0.50 | 0.45 |
| AL | >30-90 | 0.05 | 0.55 | 0.4 |
| PQ *(AL+PQ and DP+PQ)* | >30-90 | 0.05 | 0.50 | 0.45 |
| AL | ≥90 | 0.05 | 0.45 | 0.50 |
| PQ *(AL+PQ and DP+PQ)* | ≥90 | 0.05 | 0.40 | 0.55 |

**Table S8.** Prior probability distribution in Peru

| Community | Prior probability of recrudescence (C) | Prior probability of relapse (L) | Prior probability of reinfection (I) |
| --- | --- | --- | --- |
| Cahuide | 0.275 | 0.450 | 0.275 |
| Lupuna | 0.330 | 0.330 | 0.330 |

**Table S9.** Prior probability combinations for the sensitivity analysis for recurrence classification in both cohorts

| Run | Prior probability of recrudescence (C) | Prior probability of relapse (L) | Prior probability of reinfection (I) |
|---|---|---|---|
| 1 | 0.33 | 0.33 | 0.33 |
| 2 | 0.95 | 0.025 | 0.025 |
| 3 | 0.025 | 0.025 | 0.95 |
| 4 | 0.025 | 0.95 | 0.025 |
